# Financial Meltdown in “Swing Hospitals” during the COVID-19 Outbreak

**DOI:** 10.1101/2021.06.06.21258442

**Authors:** Reo Takaku, Izumi Yokoyama

## Abstract

Hospitals in Japan have complete autonomy in deciding whether to admit COVID-19 patients, similar to that of the US. Taking this into account, we estimated the effect of admitting COVID-19 patients on hospital profits, using instrumental variable (IV) regression. Using IVs related to government intervention enabled us to not only estimate the effect of admitting COVID-19 patients among “swing hospitals,” where both options (to admit or to not admit COVID-19 patients) could potentially be realized but to also evaluate the effect of government intervention on such hospitals. Our empirical results revealed that monthly profits per bed decreased by approximately JPY 600,000 (*≈* USD 6,000) among swing hospitals, which is 15 times the average monthly profits in 2019. This overwhelming financial damage indicates that it would be costly for swing hospitals to treat COVID-19 patients because of their low suitability for admitting such patients. Given the implications of our main results, we propose an alternative strategy to handling surges in patients with new infectious diseases.

## 1 Introduction

The potential financial meltdown of hospitals from the admission of COVID-19 patients is a worldwide policy issue. This problem is particularly relevant in countries such as the US and Japan, where private hospitals predominantly provide care for COVID-19 patients (American Hospital Association, 2020; Japan Hospital Association, 2020). Thus, there is an urgent need to estimate the exact effect the admitting of COVID-19 patients has on hospital finances.

More specifically, this estimation inevitably needs to be done for the following reasons: First, it is necessary to secure capacity for the treatment of COVID-19 patients by preventing private hospitals from shutting down, which can be enabled by recognizing the financial burden from accepting COVID-19 patients. Second, hospital finances can affect the quality of hospital care, and thus to keep a proper quality of service, we need to understand the costs of admitting COVID-19 patients.^1^ In fact, over 40% of Japanese medical institutions lowered their bonuses for nurses and other staff members in 2020 (Japan Times, 2020) to overcome financial difficulties, which can potentially reduce the motivation of hospital staffs and hence the quality of care. Third, obtaining some evidence on the impact of admitting COVID-19 patients can influence policymakers’ decisions since implementing a policy requires accurate information on the exact amount of financial damage and the mechanism that could cause it.

Despite the importance of estimating the impact of admitting COVID-19 patients on hospital finance, to the best of our knowledge, there has been no empirical analysis on this topic using hospital-level data. However, fortunately, we were given access to the extremely valuable data that the Tokyo Metropolitan Government collected in order to review hospitals’ financial conditions during the COVID-19 outbreak. Thus, by utilizing this unique and timely panel data on hospitals in Tokyo, in this study, we explore how a pandemic affects hospital finances.

Indeed, this study is the first econometric analysis that successfully and rigorously estimates the effect of admitting COVID-19 patients on hospital finances considering heterogeneity in suitability for admitting COVID-19 patients among hospitals.^2^

Next, in spite of how countries such as the US and Japan whose private hospitals provide care for COVID-19 patients should have faced similar situations and problems, why do we pay special attention to Japan? There are two main reasons for that: First, private hospitals in the US and Japan are allowed to choose whether to admit COVID-19 patients without any explicit penalties for refusing them (American Hospital Association, 2020; Japan Hospital Association, 2020). In the US, however, it is possible for a state governor to implement the “surge and flex” protocol and mandate hospitals to expand their bed capacity, as Andrew M. Cuomo did in New York State, while the decision is fully voluntary in Japan.^3^

In such an environment, we need to consider the self-selection behaviors of hospitals when estimating the effect of admitting COVID-19 patients on profits; for that reason, we used instrumental variable (IV) estimation. Considering that the IV regression estimates the local average treatment effect (or LATE) (Imbens and Angrist, 1994), what we estimate is equivalent to the effect among swing hospitals where both of the two options (admit or not admit COVID-19 patients) could potentially be realized. Moreover, by using IVs related to governmental intervention, we can estimate not only the effect of COVID-19 admissions among swing hospitals but also the effect of government intervention on those hospitals because their choices can be affected by the intervention. In other words, we intentionally focus on estimating the effects on swing hospitals because they are the only hospitals whose decisions can be changed by government intervention. Thus, to separate the effect on swing hospitals from effects of other types and to obtain implication for government intervention, analyzing Japan, where the degree of freedom in choosing whether to admit COVID-19 patients is higher than that in the US, should be more suitable.

Second, note that Tokyo is a unique locale and can be thought to be an ideal laboratory for estimating the effects of COVID-19 patient admission on profits. This is because a huge number of hospitals are geographically close and environmentally homogeneous in Tokyo. In fact, the hospital bed density in Tokyo is higher than that in New York City, London, and Paris, and it is probably the highest in the world (Rodwin and Gusmano, 2006), although this might be surprising to many of us. This enabled us to uncover the financial consequences of COVID-19 with great precision because we can compare many hospitals with similar environmental characteristics. In other words, without heterogeneity in environmental characteristics, it becomes easier to elicit the pure effects of COVID-19 patient admission on profits.

Utilizing these advantages in estimations, our empirical results reveal that monthly profits per bed decreased by approximately JPY 600,000 (*≈* USD 6,000) among swing hospitals^4^, 15 times the average monthly profits in 2019. Further, we also implement a detailed analysis of the hospitals’ characteristics by hospital type, which revealed the mechanism that yielded the huge losses among swing hospitals. Indeed, it was found that this huge reduction of profits is driven by the cancellation of standard medical care. Although this finding has been widely reported in the US and Japan (American Hospital Association, 2020; Japan Hospital Association, 2020; Moynihan et al, 2020; GHC, 2020), our study is the first to clearly demonstrate not only the difference in the situations of financial damages among hospital types but also the reasons why each financial situation was realized for each hospital type.

By considering hospitals’ heterogeneity in suitability for admitting COVID-19 patients, we have found important implications concerning hospital capacity policy for COVID-19 patients: The Japanese government is currently further expanding hospital capacity by requesting beds for COVID-19 patients. Our results indicate, however, that this intervention may again induce swing hospitals (“compliers”) to admit COVID-19 patients, even when these hospitals are not suitable for such admissions. By correctly understanding how each type of hospital is affected by government intervention, we will be able to avoid repeating the same disastrous experience —huge financial losses at swing hospitals. Preventing such financial losses will, in turn, prevent the abrupt closure of hospitals and contribute to securing sufficient capacity for the treatment of COVID-19 patients. It goes without saying that the implication we obtained here can be applied to any country such as the US where private hospitals provide care for COVID-19 patients.

## 2 Background

### 2.1 COVID-19 Outbreak in Tokyo

Throughout its fight against COVID-19, Japan has had the lowest number of confirmed COVID-19 infections and related deaths among G7 countries. Even in early April 2020, the number of newly confirmed cases in Tokyo did not exceed 200 per day. The number of inpatients with COVID-19 was approximately 3,000, even at the peak of the pandemic. Given that the population in Tokyo is about 14 million and that the Japanese government did not implement strict measures, these statistics seem miraculous (Aldrich and Yoshida, 2020).

However, it should be noted again that most COVID-19 patients are treated by nongovernmental hospitals in Tokyo, without any explicit support from the government. Therefore, regardless of the relatively mild spread of COVID-19 in terms of international comparison, the increase of inpatients with COVID-19 gave serious pressure on each private hospital as well as a serious threat to the continued functioning of hospitals. More details on hospital ownership and the COVID-19 outbreak in Tokyo are summarized in Online Appendix A.

### 2.2 Expanding Hospital Capacity

To deal with the surge in COVID-19 patients and strike a balance between medical care for patients with and without COVID-19, many countries have constructed so-called “temporary hospitals” for COVID-19 patients who exhibit moderate to mild symptoms.^5^ The care provided in these countries for COVID-19 patients is mainly in large hospitals. For example, in the UK, Barts Health NHS Trust, which has 1,800 beds, offered 800 beds to patients with COVID-19 at the end of January 2021 (NHS England and NHS Improvement Website, 2021).^6^

This approach has not yet been adopted in Japan. On February 9, 2020, the Ministry of Health, Labour and Welfare (MHLW) in Japan ordered local governments to expand hospital capacity for COVID-19 patients and provided brief guidelines regarding which kind of hospitals should treat patients with COVID-19. In the guidelines, the MHLW made the following three requirements for general hospitals that admit COVID-19 patients.

First, hospitals should prioritize private rooms for new COVID-positive admissions, but patients with a confirmed diagnosis can continue to be treated in the same room. Second, toilets used by COVID-positive patients should not be used by other patients. Third, hospitals should strongly adhere to the requirements for infectious diseases for a designated medical institution (i.e., institutions designated to treat patients with new infectious diseases) when admitting COVID-19 patients.^7^ As suggested in this third point, the most important requirement is that “one or more physicians with medical experience of treating infectious diseases should always be on duty.” When it comes to the case of COVID-19, in which patients exhibit symptoms similar to pneumonia, this requirement is naturally interpreted as “one or more physicians with medical experience in treating infectious ‘respiratory’ diseases should always be on duty.”

Note that these requirements from the government do not take into account non-negligible costs related to admitting COVID-19 patients, namely cancellation costs of usual medical care, the fact of which consequently led to the disastrous financial losses of swing hospitals.

Although each hospital can freely choose whether to admit COVID-19 patients, these guidelines have indeed affected some hospitals’ decisions. Further, the local government’s request to hospitals to serve COVID-19 patients influences hospitals’ decision-making to some degree. In response to that request, each hospital decides whether to accept COVID-19 patients based on its resources and the content of the guidelines provided by the MHLW.

Eventually, 95 out of 638 hospitals in Tokyo offered a total of 2,980 beds by August 2020.^8^ Although many hospitals have a large number of beds, the number of beds for COVID-19 patients per hospital was extremely small. For example, University of Tokyo hospitals only offered 30 beds, out of about 1,200 beds for COVID-19 patients.^9^ The strongest feature of Japan’s handling of the surge in COVID-19 patients was that care was shared by many hospitals, without concentrating those patients in large hospitals only.

## 3 Research Design

In this section, we first present the ordinary least squares (OLS) model, without considering the self-selection of each hospital, as a baseline model. Next, we present our identification strategy, which overcomes the endogeneity problem in the OLS model by employing the IV estimation.

### 3.1 Baseline Model

In the actual implementation of the regression analysis, we begin by estimating the following OLS as a baseline model:

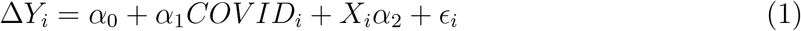

where *Y_i_* is the outcome variable in hospital *i*, such as profit per bed. Since we are interested in the change in the outcome variable during the first wave of COVID-19, we use the outcome that captures the change from February 2020 to April and May 2020. In the implementation, we first calculate the average of *Y* in April and May 2020, 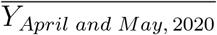, because the effect on hospitals was severe in these two months. Second, to capture the change from before to after COVID-19 outbreak, we subtract *Y_Feb,_* _2020_ from 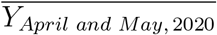. In addition to this, even without the effect of COVID-19, there should be usually some seasonal trend from February to April and May, which can be obtained from the data from the previous year. Thus, we subtract the seasonal trend captured by 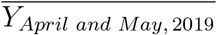− *Y_Feb,_* _2019_ from 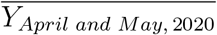− *Y_Feb,_* _2020_. Then, the outcome variable *Y* that we use in our regressions can be written as (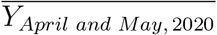− *Y_Feb,_* _2020_) − (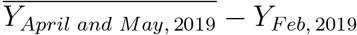 *− Y_Feb,_* _2019_).

Note that this expression can also be interpreted as the change in outcome variables during the COVID-19 outbreak, controlled for hospital fixed effects because the first difference is taken to the outcomes in each month. This interpretation is possible because we use panel data.^10^

The independent variable *COVID* is a dummy variable that takes one if hospital *i* have admitted COVID-19 patients and zero otherwise. *X_i_* is a vector of predetermined covariates, such as the local prevalence of COVID-19. We call this variable “*Case*.” To evaluate the local prevalence of COVID-19, we construct the average number of newly confirmed COVID-19 cases around each hospital in April and May. More concretely, using the exact address of all hospitals in Tokyo, we measure the distance from the city center to each hospital and use it as a weight to construct *Case*.^11^

Note that this OLS model does not consider the fact that each hospital can decide whether to admit COVID-19 patients, which makes the status of admitting COVID-19 patients endogenous. Thus, it is expected that the OLS estimate of the coefficient of *COVID* suffers from bias by ignoring the endogeneity. For example, hospitals expecting improved finances from admitting COVID-19 patients might be likely to do so, if their decision is made based on comparing costs and benefits. In this case, the endogenous decision will upwardly bias the estimated effects on profits in an OLS analysis.

### 3.2 Identification

To consider the endogenous decision-making of admitting COVID-19 patients, we implement IV regressions. Note that ideal IVs strictly satisfy the conditions of relevancy and exclusion restriction. To satisfy both conditions, we use two of the characteristics stated in the national government guidelines as our IVs. In Section 2.2, we already confirmed that the relevancy condition was likely to be satisfied, that is, the guidelines’ requirements affect hospitals’ decision-making, which in turn satisfies the condition of relevancy.

In contrast, to satisfy the condition of exclusion restriction, IVs must affect hospital finances only through COVID-19 patient admission. The guidelines did not take into account the potential detrimental effects on hospital finances of admitting COVID-19 patients, because they were issued in early February when the number of COVID-19 cases was low. Therefore, the guidelines can be regarded as an exogenous shock to hospitals and their financial situations.

Fortunately, the characteristics of hospitals described in the guidelines are an ideal set of IVs (MHLW, 2020b), thus we exploit characteristics of hospitals as IVs. Our first IV is the number of respiratory specialists per bed. This corresponds to the third requirement presented in Section 2.2. Because COVID-19 is a respiratory disease, it is difficult for hospitals without respiratory specialists to admit patients even if they want to. Importantly, since respiratory care only accounts for a small part of the medical revenue of average hospitals, it is reasonable to suppose that respiratory specialists affect hospital finances only through the admission of COVID-19 patients.

Our second IV is the number of private rooms per bed, corresponding to the first requirement in Section 2.2. Since COVID-19 is a highly infectious disease, even in early February, the MHLW recommended that patients stay in private rooms (MHLW, 2020b). This recommendation was strictly enforced during the first outbreak in Tokyo. The number of private rooms per bed may not have a large influence on hospital finances normally, but it is strongly associated with the admission of COVID-19 patients; therefore, it is expected to work well as an IV.

Lastly, the local prevalence of COVID-19 could directly affect the financial outcomes of hospitals. In addition to this, especially in areas where local prevalence of COVID-19 is high, it might be possible that the fact that there are respiratory specialists in a hospital can prevent people from going to the hospital because they may expect that the hospital can handle patients suspected of being infected with COVID-19 as well and hence the risk of infection inside the hospital is high. Then, whether there are respiratory specialists in a hospital could affect hospital finances by the decrease in the number of potential non-COVID-19 outpatients, regardless of whether the hospitals admit COVID-19 patients, and this could occur especially in hospitals near epidemic areas. To consider this possibility, although we already use an ideal set of IVs, we control for the local prevalence of COVID-19, *Case*, to be more robust against violation of the exclusion restriction.

### 3.3 IV Regression

To uncover the effects of admitting COVID-19 patients on financial variables after considering the self-selection behavior of each hospital, we implement IV regressions using the IVs presented in the previous section.

First, we will check that the IVs are sufficiently correlated with the admission status (*COV ID_i_*) in the first stage, as follows:

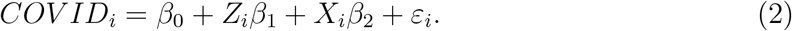

where *Z_i_* is a vector of the instrument variables and *X_i_* is an exogenous variable common to that used in the second-stage regression, i.e., the local prevalence of COVID-19, “*Case*.”^12^ *ε_i_* denotes the error term. In this specification, *β*_1_ captures the effect of each IV on the admission of COVID-19 patients.

Next, the second-stage regression is estimated using the predicted value of *COV ID_i_* obtained from the first-stage regression, as follows:

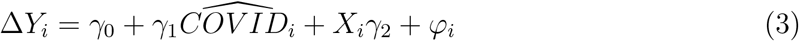

where 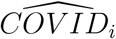 is *COV ID_i_* instrumented by the IVs and the exogenous variable *X_i_*. As explained in Section 3.2, to be more robust against violation of the exclusion restriction, we control for the local prevalence of COVID-19 as *X_i_*, which is represented by “*Case*.” In this equation, *γ*_1_ is interpreted as the LATE, which measures the effect of COVID-19 patient admission among compliers, i.e., swing hospitals, hospitals in which the admission status can be changed by the instruments. Finally, standard errors clustered at the 12 medical areas were reported.

## 4 Data and Descriptive Statistics

### 4.1 Data

The data used in this study were collected by the Tokyo Metropolitan Government in July 2020 to uncover the financial effects of the COVID-19 outbreak on hospitals. The survey was sent to all hospitals in Tokyo (N = 642), and we received 332 responses (response rate = 51.7 %). The data cover the period from February to May 2020. We also surveyed all variables in the same months in 2019 in order to compare hospitals’ pre- and post-COVID-19 financial situations. The data include a variety of financial variables such as total medical revenue and total costs, as well as the number of patients, but do not include information on the number of physicians by specialty.

Thus, to compensate for the lack of information on the number of physicians by specialty, we used the physician data compiled by Nihon Ultmarc as of October 2017 (Takaku, 2020).^13^ Using this database, we can know the number of respiratory and other specialists for the period *before* the COVID-19 outbreak, which is necessary for our study because the number of respiratory specialists per bed is one of our IVs.

Further, we excluded the data of 50 hospitals for the following two reasons. First, some specialty hospitals will never be able to accept COVID-19 patients and, thus, have no option to accept COVID-19 patients, in which case, there is no self-selection behavior among hospitals. For example, 36 hospitals are psychiatric hospitals and six are for rehabilitation; some other hospitals provide only dental care or care for children with special needs and, thus, were excluded from our analysis. Second, 40 public hospitals were excluded because their financial structure is completely different from that of private hospitals. After eliminating some missing values, the sample size in the main analysis is 222 general private hospitals.

### 4.2 Descriptive Statistics

The descriptive statistics of our main analysis are shown in Columns 1 to 3 in Table 1. The sample size in each column in Table 1 indicates that 68 hospitals admitted COVID-19 patients during the first outbreak, while 154 hospitals did not. It is striking that the treatment group (hospitals that admitted COVID-19 patients) experienced a reduction in profit per bed by JPY 358,600 (*≈* USD 3,586) in April–May on average, while the reduction in profit per bed among the control group is only JPY 161,500 (*≈* USD 1,615). We also observe a large difference in the reduction in total revenue per bed between the treatment and control groups. According to Table 1, the average number of beds *before* COVID-19 outbreak for the treatment group is 363, but only 137 for the control group. This suggests that large hospitals are more likely to admit COVID-19 patients. The number of newly confirmed cases around hospitals is also larger in the treatment group.

**Table 1:**
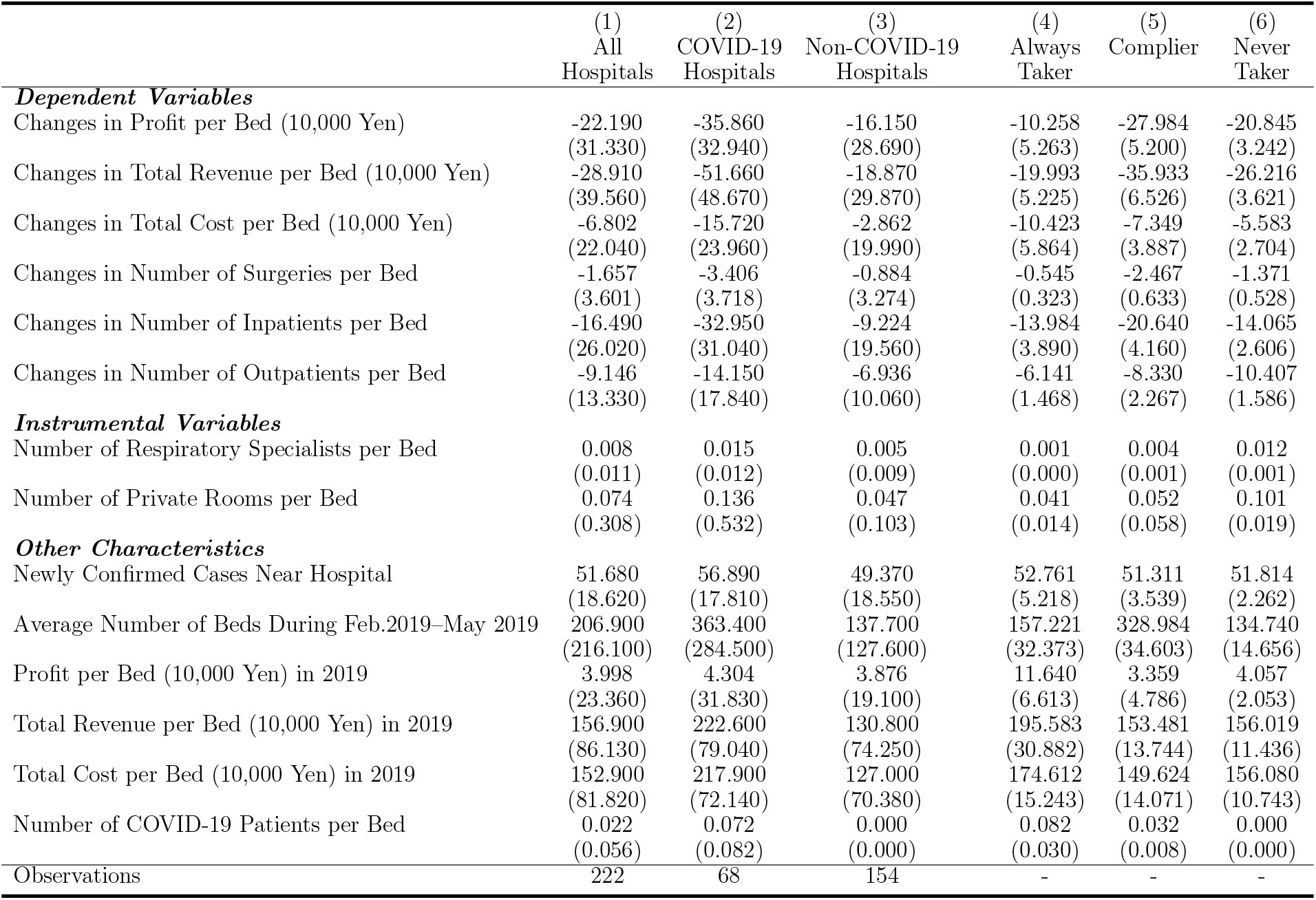
Descriptive Statistics *Notes:* Standard deviations are in parentheses in Columns 1 to 3. Bootstrap standard errors are in parentheses in Columns 4 to 6. The method to derive summary statistics for always-takers, compliers, and never-takers is explained in Online Appendix C.

|  | (1)<br>All<br>Hospitals | (2)<br>COVID-19<br>Hospitals | (3)<br>Non-COVID-19<br>Hospitals | (4)<br>Always<br>Taker | (5)<br>Complier | (6)<br>Never<br>Taker |
| --- | --- | --- | --- | --- | --- | --- |
| ***Dependent Variables*** |  |  |  |  |  |  |
| Changes in Profit per Bed (10,000 Yen) | -22.190<br>(31.330) | -35.860<br>(32.940) | -16.150<br>(28.690) | -10.258<br>(5.263) | -27.984<br>(5.200) | -20.845<br>(3.242) |
| Changes in Total Revenue per Bed (10,000 Yen) | -28.910<br>(39.560) | -51.660<br>(48.670) | -18.870<br>(29.870) | -19.993<br>(5.225) | -35.933<br>(6.526) | -26.216<br>(3.621) |
| Changes in Total Cost per Bed (10,000 Yen) | -6.802<br>(22.040) | -15.720<br>(23.960) | -2.862<br>(19.990) | -10.423<br>(5.864) | -7.349<br>(3.887) | -5.583<br>(2.704) |
| Changes in Number of Surgeries per Bed | -1.657<br>(3.601) | -3.406<br>(3.718) | -0.884<br>(3.274) | -0.545<br>(0.323) | -2.467<br>(0.633) | -1.371<br>(0.528) |
| Changes in Number of Inpatients per Bed | -16.490<br>(26.020) | -32.950<br>(31.040) | -9.224<br>(19.560) | -13.984<br>(3.890) | -20.640<br>(4.160) | -14.065<br>(2.606) |
| Changes in Number of Outpatients per Bed | -9.146<br>(13.330) | -14.150<br>(17.840) | -6.936<br>(10.060) | -6.141<br>(1.468) | -8.330<br>(2.267) | -10.407<br>(1.586) |
| ***Instrumental Variables*** |  |  |  |  |  |  |
| Number of Respiratory Specialists per Bed | 0.008<br>(0.011) | 0.015<br>(0.012) | 0.005<br>(0.009) | 0.001<br>(0.000) | 0.004<br>(0.001) | 0.012<br>(0.001) |
| Number of Private Rooms per Bed | 0.074<br>(0.308) | 0.136<br>(0.532) | 0.047<br>(0.103) | 0.041<br>(0.014) | 0.052<br>(0.058) | 0.101<br>(0.019) |
| ***Other Characteristics*** |  |  |  |  |  |  |
| Newly Confirmed Cases Near Hospital | 51.680<br>(18.620) | 56.890<br>(17.810) | 49.370<br>(18.550) | 52.761<br>(5.218) | 51.311<br>(3.539) | 51.814<br>(2.262) |
| Average Number of Beds During Feb.2019–May 2019 | 206.900<br>(216.100) | 363.400<br>(284.500) | 137.700<br>(127.600) | 157.221<br>(32.373) | 328.984<br>(34.603) | 134.740<br>(14.656) |
| Profit per Bed (10,000 Yen) in 2019 | 3.998<br>(23.360) | 4.304<br>(31.830) | 3.876<br>(19.100) | 11.640<br>(6.613) | 3.359<br>(4.786) | 4.057<br>(2.053) |
| Total Revenue per Bed (10,000 Yen) in 2019 | 156.900<br>(86.130) | 222.600<br>(79.040) | 130.800<br>(74.250) | 195.583<br>(30.882) | 153.481<br>(13.744) | 156.019<br>(11.436) |
| Total Cost per Bed (10,000 Yen) in 2019 | 152.900<br>(81.820) | 217.900<br>(72.140) | 127.000<br>(70.380) | 174.612<br>(15.243) | 149.624<br>(14.071) | 156.080<br>(10.743) |
| Number of COVID-19 Patients per Bed | 0.022<br>(0.056) | 0.072<br>(0.082) | 0.000<br>(0.000) | 0.082<br>(0.030) | 0.032<br>(0.008) | 0.000<br>(0.000) |
| Observations | 222 | 68 | 154 | - | - | - |

Further, we have also carefully checked the external validity of our findings. Needless to say, in a sense that both in the US and Japan, private hospitals predominantly provide care for COVID-19 patients, we have already high external validity toward the US because of the similar systems. In addition to this, we also compare the average number of beds per hospital among OECD countries because the number of beds is the most representative and widely cited characteristic of hospitals, and it can be compared across countries with high accuracy. According to Figure A1, the average number of beds per hospital in our data is very close to the average of OECD countries. Thus, while healthcare systems greatly differ across countries, the size of hospitals in our data is very common in the OECD countries, which implies that we do not pick up specific hospitals.

### 4.3 Complier Characteristics

The IV estimate represents the LATE, which is the average treatment effect among compliers. Thus, it is extremely important to understand the characteristics of compliers. To this end, we estimate the average characteristics of compliers in comparison with those of always-takers and never-takers, employing the method used in the following previous studies (Kowalski, 2016; Abrigo et al, 2019; Marbach and Hangartner, 2020). blueIn this process, we transform our two continuous IVs in our most preferred specification into one binary indicator using principal component analysis (for more details, see Online Appendix C).^14^ Our results are shown in Columns 4 to 6 in Table 1.

If we compare the predetermined variables among the three groups, the most notable difference between compliers and other types is hospital size. The average number of beds for always-takers is 157 and 134 for never-takers. In contrast, the average number of beds for compliers is 328, suggesting that the size of the hospitals among compliers, in terms of the number of beds, is more than twice that of always- and never-takers.

Despite the hospital size that is much smaller than that of compliers, the revenue per bed in 2019 was larger for always-takers than for compliers—JPY 1,955,830 (*≈* USD 19,558.3) for always-takers. This is approximately 25% more than the revenue of compliers. Note that there is no such large difference in the total costs in 2019 between the two types, which implies low profitability among compliers; this is consistent with the average amount of profit per bed in 2019. In sum, compliers are originally relatively large hospitals with low profitability. By contrast, it turned out that always-takers admitted COVID-19 patients despite their smaller size and fewer respiratory specialists and private rooms per bed, compared to the characteristics of compliers.

Thus, it should have been very difficult, especially for compliers (mostly large hospitals), to implement effective zoning because they usually treat a large number of patients with a variety of diseases. The low profitability among compliers even in the period *before* the COVID-19 outbreak also implies that they had originally admitted many patients with various diseases rather than having specialized in a limited number of diseases. As a result, compliers should have had to cancel much more general medical care than always-takers should have.

Next, we will confirm the changes in outcome variables from *before* to *after* COVID-19 outbreak. The average changes in the number of surgeries per bed and inpatients, whose values are presented in the category “Dependent Variable” in Table 1, provide strong evidence for large-scale cancellation of standard medical care among compliers. Especially, the decrease in the number of inpatients per bed among compliers is much larger than that among always-takers.

Finally, despite the higher level of the cancellation of standard medical care, the number of COVID-19 patients per bed for compliers is lower (0.032) than that for always takers (0.082). This again is consistent with the fact that compliers had been “swinging” between the two options of whether to admit COVID-19 patients or not, while always-takers had originally decided to admit such patients. This difference in their behaviors should reflect the fact that compliers are mostly “unsuitable” hospitals in admitting COVID-19 patients.

## 5 Regression Results

### 5.1 First-Stage Regression Results

Columns 1 and 2 in Table 2 report the results of the first-stage regression of the IV estimations when *COV ID_i_* is regressed on our IV(s) only, while Columns 3 and 4 report the results when *Cases* is also controlled for.

**Table 2:** First-stage Regression *Notes:* The dependent variable, *COVID*, is a dummy variable that takes one if each hospital has admitted COVID-19 patients and zero otherwise. Standard errors clustered at the level of 12 medical areas are reported in parentheses. *Cases* is the monthly number of COVID-19 patients around each hospital. *** p<0.01; ** p<0.05; * p<0.1.

| Dependent Variable: COVID | (1) | (2) | (3) | (4) |
| --- | --- | --- | --- | --- |
| Number of Respiratory Specialists per Bed | 18.361***<br>(1.571) | 18.113***<br>(1.555) | 17.421***<br>(1.535) | 17.079***<br>(1.536) |
| Number of Private Rooms per Bed | - | 0.170***<br>(0.030) | - | 0.182***<br>(0.032) |
| Cases | - | - | 0.003<br>(0.002) | 0.003*<br>(0.002) |
| Constant | 0.167***<br>(0.017) | 0.156***<br>(0.017) | 0.042<br>(0.081) | 0.020<br>(0.076) |
| R-squared | 0.177 | 0.190 | 0.187 | 0.202 |
| Mean of Dependent Variable | 0.306 | 0.306 | 0.306 | 0.306 |
| Observations | 222 | 222 | 222 | 222 |
| **F Test ($H_0$: Coefficient(s) of IV(s)=0)** |  |  |  |  |
| F-statistics | 136.678 | 155.132 | 128.887 | 109.458 |
| P-value | 0.000 | 0.000 | 0.000 | 0.000 |

According to Table 2, both IVs, the number of respiratory specialists per bed and number of private rooms per bed, correlate positively with the probability of hospitals admitting COVID-19 patients even at the 1% significance level in all columns. First, note that the positive correlation between the IVs and *COVID* is exactly what we have expected in Section 3.2. Furthermore, the F-statistics from the F-test are also very large enough to pass the F-test, which indicates that the IVs strongly satisfy the IV relevancy condition. Thus, we will move on to the second-stage regression (SSR).

### 5.2 Second-Stage Regression Results

Table 3 reports the main results of the SSR on profits. The results of OLS appear in Columns (1) and (2), without and with *Cases* controlled for, respectively. Columns (3) to (6) show the results of the IV regressions. Columns (3)-(4) and Columns (5)-(6) report the results without and with *Cases* controlled for, respectively. As explained in Section 3.2, controlling for *Cases* is more likely to satisfy the exclusion restriction.

**Table 3:** Main Results on Profit *Notes:* The dependent variable is the year-on-year differences in each variable, divided by the number of beds. The unit is 10,000 JPY. *Cases* is the monthly number of COVID-19 patients around each hospital. Standard errors clustered at the level of 12 medical areas are reported in parentheses. *** p<0.01; ** p<0.05; * p<0.1.

|  | (1) | (2) | (3) | (4) | (5) | (6) |
| --- | --- | --- | --- | --- | --- | --- |
| Dependent Variable: $Y = (\overline{y_{April\ and\ May, 2020}} - y_{Feb, 2020}) - (\overline{y_{April\ and\ May, 2019}} - y_{Feb, 2019})$ | | | | | | |
| $y$: Profit | | | | | | |
| Estimation Method: | OLS |  | IV |  |  |  |
| COVID | -19.706***<br>(4.758) | -17.156***<br>(4.495) | -62.078***<br>(9.957) | -60.613***<br>(9.621) | -59.193***<br>(12.216) | -57.939***<br>(11.635) |
| Cases | - | -0.339***<br>(0.105) | - | - | -0.145<br>(0.139) | -0.151<br>(0.138) |
| Constant | -16.154***<br>(4.133) | 0.602<br>(2.657) | -3.175<br>(4.504) | -3.623<br>(4.391) | 3.422<br>(3.048) | 3.338<br>(2.918) |
| R-squared | 0.084 | 0.124 | - | - | - | - |
| Mean of Y With COVID=0 | -16.154 | -16.154 | -16.154 | -16.154 | -16.154 | -16.154 |
| Observations | 222 | 222 | 222 | 222 | 222 | 222 |
| **Instrumental Variables** |  |  |  |  |  |  |
| Number of Respiratory Specialists | - | - | Yes | Yes | Yes | Yes |
| Number of Private Rooms | - | - | No | Yes | No | Yes |
| **Tests of Overidentifying Restrictions ($H_0$: All IVs Exogenous)** |  |  |  |  |  |  |
| Basmann Chi-squared Statistic | - | - | - | 0.209 | - | 0.106 |
| P-value | - | - | - | 0.647 | - | 0.744 |
| Sargan Chi-squared Statistic | - | - | - | 0.212 | - | 0.108 |
| P-value | - | - | - | 0.645 | - | 0.742 |

Concerning the OLS results, note that the dependent variable defined in the table is conceptually equivalent to the graphical representation in Figure B(a) in Online Appendix B because we use the adjusted and de-trended outcomes in the regression model in Table 3.

According to Columns (1) and (2), regardless of controlling for *Case*, the OLS estimates on the coefficient of *COVID* are significantly negative even at the 1% significance level, and the coefficients suggest that the monthly profits decreased by approximately JPY 170,000 to 190,000 (*≈*USD 1,700 to 1,900) per bed. However, we should not forget that the OLS estimates do not consider the self-selection behavior of hospitals.

In the IV regressions, the estimated coefficients of *COVID* are much smaller (i.e., larger negative) than in the OLS regression. It is, of course, possible to understand the difference between the OLS and IV estimates as being consistent with the upward bias suggested in Section 3.1. In addition to this potential bias in the OLS caused by ignoring the self-selection behaviors among hospitals, the difference between the OLS estimate and the IV estimate can also stem from the fact that the IV estimates are the LATE, i.e., effects among compliers only. Thus, the difference between the OLS and IV estimates in Table 3 is also consistent with the fact that the IV estimates reflect only the extremely large damage to the profits among compliers, while the OLS estimates also include the relatively small damage among always-takers.

In Column (6), we use two instrumental variables and control for the local prevalence of COVID-19, which is the most preferred specification in this main analysis. In this specification, the coefficient on *COVID* is -57.939 and significantly negative even at the 1% significance level. The monthly profits decreased by approximately 600,000 JPY (*≈* USD 6,000) per bed, and this result is stable across specifications as well as the choice of IVs. Surprisingly, the amount of JPY 600,000 is 15 times the average monthly profits in 2019 (JPY 39,980) according to Table 1.

The coefficients estimated by the IV estimation are robust and stable across specifications and IV choices, which suggests that the exclusion restriction is satisfied for all the IVs. In other words, if even one IV was not valid in terms of the exclusion restriction, we should have obtained more unstable IV estimates over the choice of IVs. The robustness of the exclusion restriction can also be confirmed by the over-identifying restrictions tests, the results of which strongly suggest that all IVs are exogenous.

### 5.3 Results on Other Outcomes

Next, to explore the mechanisms behind this large deterioration in profits, we also report the results for other outcome variables in Figure 1, and the detailed regression table is presented in Online Appendix D.

**Figure 1:**
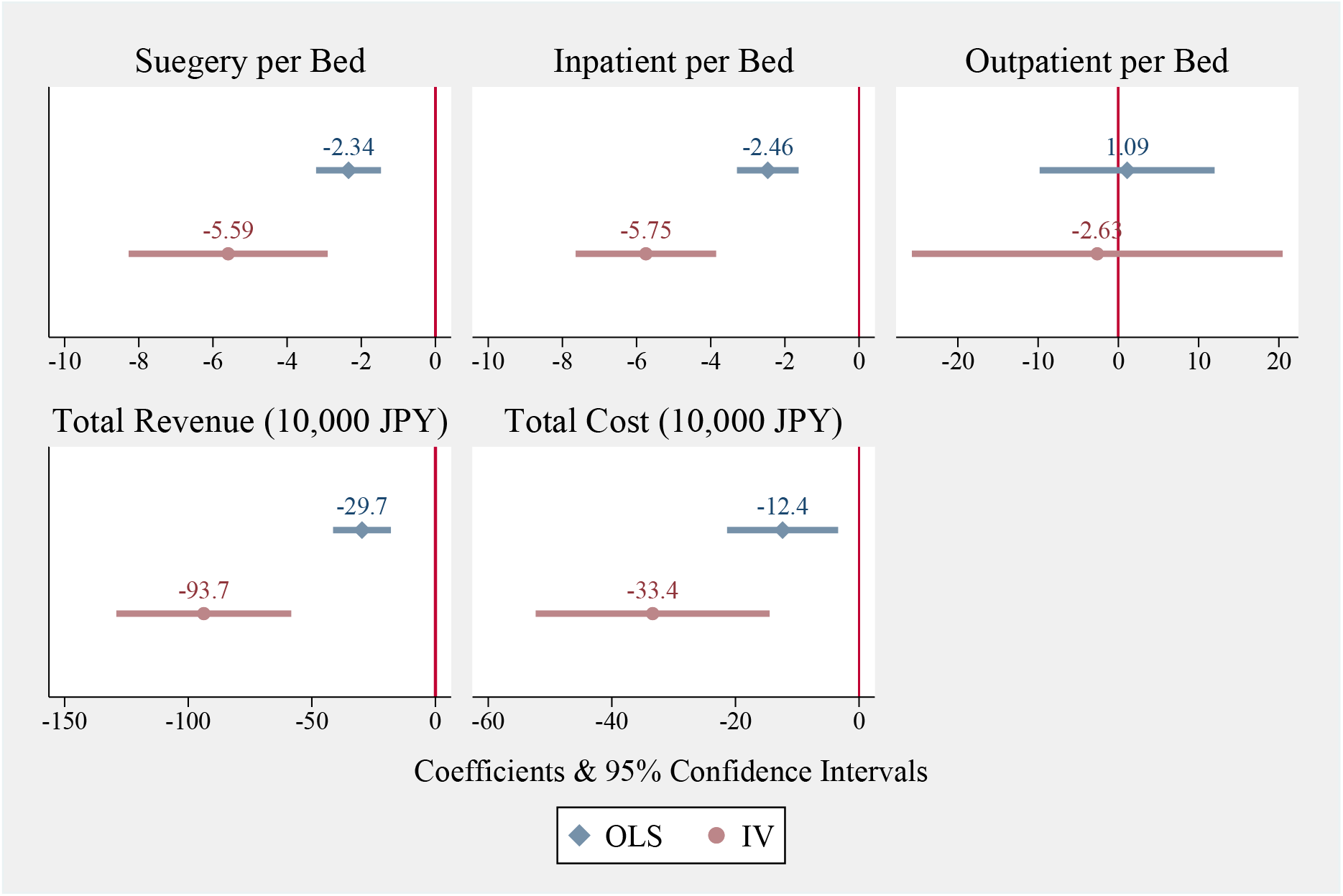
Results on Other Outcomes *Notes:* The vertical line represents 0. The estimated coefficients and 95 percent confidence intervals of *COVID* from both OLS and IV estimation are shown in each figure.The regression table that the results of this figure based on is presented in Online Appendix D. The regression specification of OLS and IV is the same as Columns (2) and (6) in Table 3, respectively.

From the IV estimates in Figure 1, we can see that the reduction in profits was driven by the sharp reduction in the number of inpatients as well as the number of surgeries. In addition to this, we find the magnitude of the IV estimates far larger than that of the OLS counterparts in these outcomes.

Finally, IV results on the revenue per bed reveal the reduction of JPY 937,000 among compliers. These results again suggest that the cancellation of usual medical care is a non-negligible cost of admitting COVID-19 patients (American Hospital As- sociation, 2020; GHC, 2020). Note that our supplemental results in Online Appendix D and E also support the view that the main costs of admitting COVID-19 patients arise from the cancellation of the usual medical care, rather than from direct medical costs.

### 5.4 Checks for Identifying Assumptions

To confirm the validity of our IVs, we implement two placebo tests. First, we use the number of physicians with other specialties per bed as a fake IV for the number of respiratory physicians per bed. This type of placebo test is useful to check for the possibility of violation of the exclusion restriction. For example, if the number of respiratory physicians was associated with the trend in profits not only through admitting COVID-19 patients but also through other channels, it is likely that the number of physicians with other specialties would also be associated with the trend in profits (i.e., outcome) and the decision whether to admit COVID-19 patients (i.e., the endogenous variable). The results of the placebo tests are shown in Figure 2. In this figure, we show the coefficient and 95% confidence intervals of the number of physicians with various specialties, including respiratory diseases (i.e., the real IV). Here, for both the first-stage and reduced-form regressions, we found the largest coefficient for the real IV, namely, the number of respiratory physicians per bed. Some of the fake IVs are also statistically significant in the first-stage and reduced-form regressions, which could happen as long as they were correlated with the number of respiratory physicians and other hospital-level unobservables. However, the point estimates of those fake IVs are very small even if these are statistically significant. Therefore, we can reasonably conclude that our real IV (the number of respiratory physicians per bed) is associated with the outcome variables only because of the admission of COVID-19 patients.

**Figure 2:**
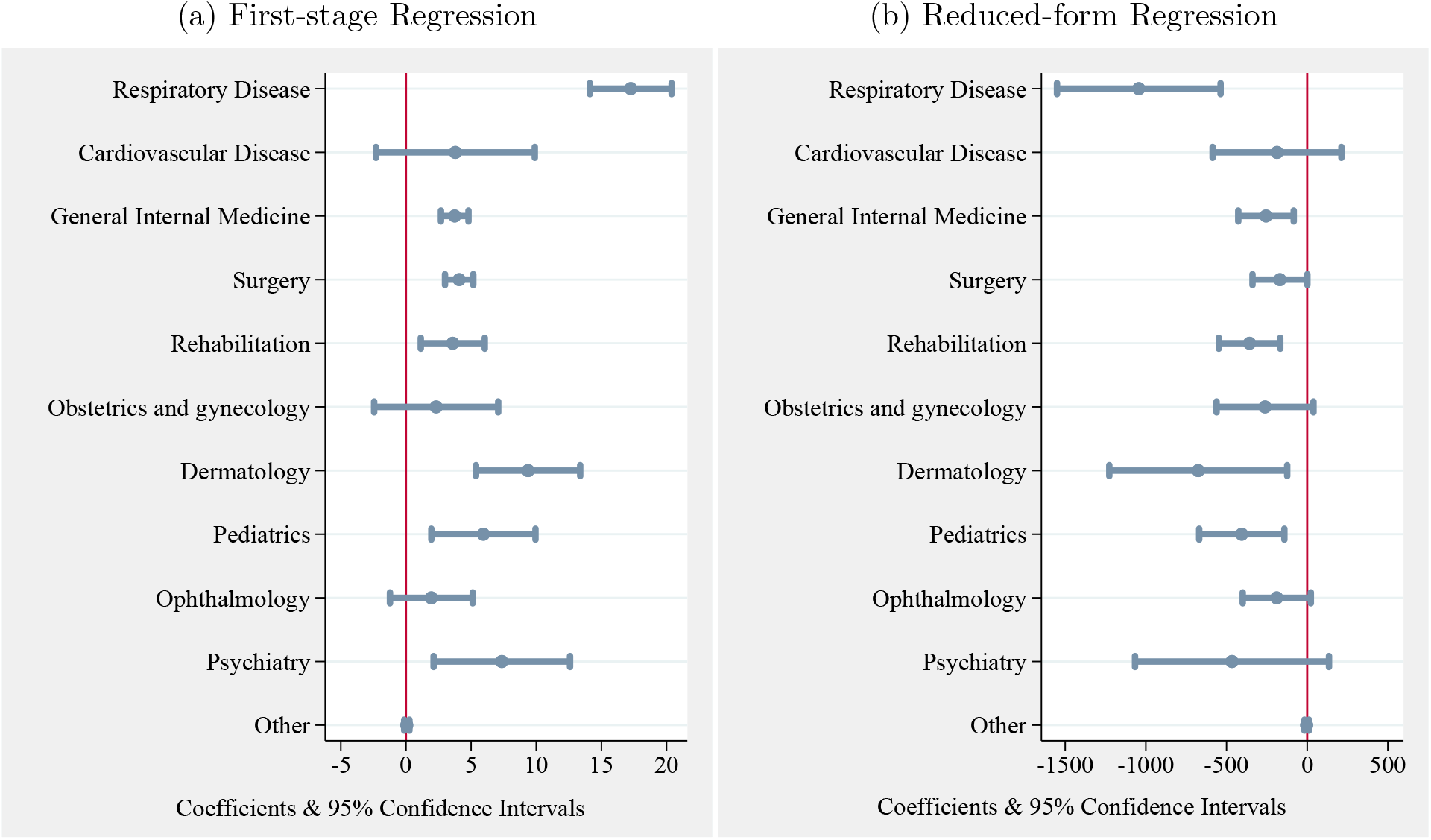
Placebo Test *Notes:* In the first-stage regressions, we estimate Equation 2 with the number of physicians of various specialties as fake instruments, after controlling for the local prevalence of COVID-19 (*Case_i_*). In the reduced-form regression, we estimate the same regression using Equation 2 by replacing the outcome variable with 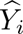. The number of physicians was standardized by the number of beds in all estimations. The unit of the outcome variable in the reduced-form regression is JPY 10,000.

Next, we have checked how our IVs were associated with the trend in profits during the period before the COVID-19 outbreak (see Online Appendix F). The results suggest that both our IVs were not associated with year-to-year changes in profits in February 2020 when COVID-19 had not yet spread throughout Japan. Again, this suggests that our IVs have no explanatory power for the trend in profits in the absence of COVID-19, but still have a strong influence on the admission of COVID-19 patients. These results reinforce the validity and reliability of our IVs.

### 5.5 Robustness Check

As shown in many epidemiological studies, the care burden for COVID-19 patients with severe symptoms is totally different from that for patients with mild symptoms.^15^ For patients with severe symptoms, hospital treatment regimens switch to costly hightech treatments, such as extracorporeal membrane oxygenation. The reimbursement for patients with severe symptoms is also very high. Therefore, we also check the robustness of our results by excluding 32 hospitals that admitted COVID-19 patients in their ICU (see Online Appendix E). The results of this subsample analysis support our main finding, that is, the strong negative effects of admitting COVID-19 patients among compliers is preserved even when we exclude hospitals that provide care for patients with severe symptoms.

Finally, although we have already discussed and confirmed the external validity for our data, we also evaluate the external validity of our findings here. More concretely, We implement supplemental analyses based on the marginal treatment effects approach Heckman and Vytlacil (2005, 2007) (see Online Appendix G). The results indicate no strong selection based on unobservable resistance to treatment, suggesting that our results have relatively high external validity in terms of unobservable characteristics that determine selection for treatment.

## 6 Concluding Remarks

In this study, we employed unique and timely monthly longitudinal data on hospital finances in Tokyo and explored how a pandemic deteriorates hospital finances considering heterogeneity in suitability for admitting COVID-19 patients among hospitals.

By exploiting the homogeneous environment in the hospital sector in Tokyo and using IV analysis to incorporate the endogenous decision of each hospital to admit COVID-19 patients, we found that admitting COVID-19 patients severely deteriorated hospital finances, especially among compliers. More specifically, our main IV estimate revealed that admitting COVID-19 patients lowered monthly profits by about JPY 600,000, which is 15 times the average monthly profits in the previous year. This IV estimate is about three times the OLS estimate, which does not consider self-selection behaviors.

To understand hospitals’ heterogeneity and their self-selection behaviors in more detail, we examined the characteristics of always-takers and compliers explicitly (Heckman and Vytlacil, 2001; Kowalski, 2016; Abrigo et al, 2019). Our analysis uncovered that compliers are relatively large hospitals with lower profits compared to always-takers. Due to the difference in characteristics, compliers had to cancel their usual provision of medical care to inpatients more frequently than always-takers because of their ineffective zoning.

All the findings in this study have an important implication for hospital capacity policy in handling the surge of COVID-19 patients. Whether the simultaneous provision of other standard care is possible or not for each hospital (having admitted COVID-19 patients) depends on factors that cannot immediately be changed, such as architectural structure and the number of medical staff. Therefore, it is extremely important to prevent hospitals with factors that can be a great disadvantage in admitting COVID-19 patients from offering beds to COVID-19 patients because simultaneously continuing their usual medical care is difficult for them. In fact, the Japanese government is currently expanding hospital capacity by requesting hospitals to offer beds for COVID-19 patients. However, as we have shown, this intervention will induce compliers to admit COVID-19 patients, although the request is not mandatory.^16^

Therefore, it is necessary to admit COVID-19 patients predominantly to large hospitals and encourage other hospitals to continue their usual medical care, as was done in the UK and other countries. We recommend avoiding missteps from the earlier phase of the pandemic in order to prevent large financial losses among compliers. Following the right path could prevent these hospitals from shutting down, thus securing sufficient capacity for the treatment of COVID-19 patients as well.

Inevitably, medical resources (e.g., specialists) across the county need to be reassigned to leading hospitals when new infectious diseases such as COVID-19 spread. However, concentrating patients in a large hospital during an emergency is incompatible with medical staff’s right to choose their workplace. In fact, it is impossible without staff members’ solidarity and understanding. Therefore, policymakers need to have a nationwide discussion about the degree of freedom medical staff should have in an emergency.

Although the number of COVID-19 cases in Japan was the lowest among G7 countries, hospitals in Japan were, ironically, on the verge of a financial meltdown. Overall, these policy implications from Tokyo are extremely important for future policies related to a pandemic from an unknown infectious disease, especially in countries with a large private health care sector, such as the US. Such countries should adopt different hospital-capacity policies for dealing with future pandemics.

## Data Availability

The data are prohibited to share anyone.

## Online Appendix

## A Background Details

### A.1 Hospitals in Japan and Tokyo

Japan has the highest number of hospitals as a ratio to the population among OECD countries (OECD, 2020). Most hospitals are small family enterprises that evolved from physicians’ offices. Large hospitals are owned by national or local governments, volunteer organizations, and universities (Ikegami and Campbell, 1995). The proportion of hospital beds owned by the government is only 22% (MHLW, 2018). Other beds are owned by non-profit organizations, such as the Japanese Red Cross and private hospitals. Non-governmental provision is an important feature of Japanese hospital care, but it should be noted that for-profit, investor-owned hospitals are prohibited.

In Tokyo, the number of hospitals is approximately 650, and the number of hospital beds is about 127,000. The proportion of beds owned by non-government hospitals is 85% (Tokyo Metropolitan Government, 2018). Given that the management of non-governmental hospitals is not sustained by general tax revenue, the COVID-19 outbreak is expected to have a particularly serious negative effect in Tokyo.

Patients can freely choose hospitals to receive treatment. Thus, many patients across the nation seek advanced treatment in famous hospitals in Tokyo. In fact, 13.4% of the hospital patients in Tokyo come from other prefectures (Tokyo Metropolitan Government, 2019). During the COVID-19 outbreak, this large inflow of patients into Tokyo suddenly stopped, as Tokyo was the epicenter of the pandemic in Japan.

In terms of hospital size, the average number of beds per hospital is 206 in our data and 197 in Japan, according to the data from OECD Health Statistics in Figure A1. These numbers are very close to the average of OECD countries. While healthcare systems greatly differ across countries, the size of hospitals in our data is very common in OECD countries.

**Figure A1:**
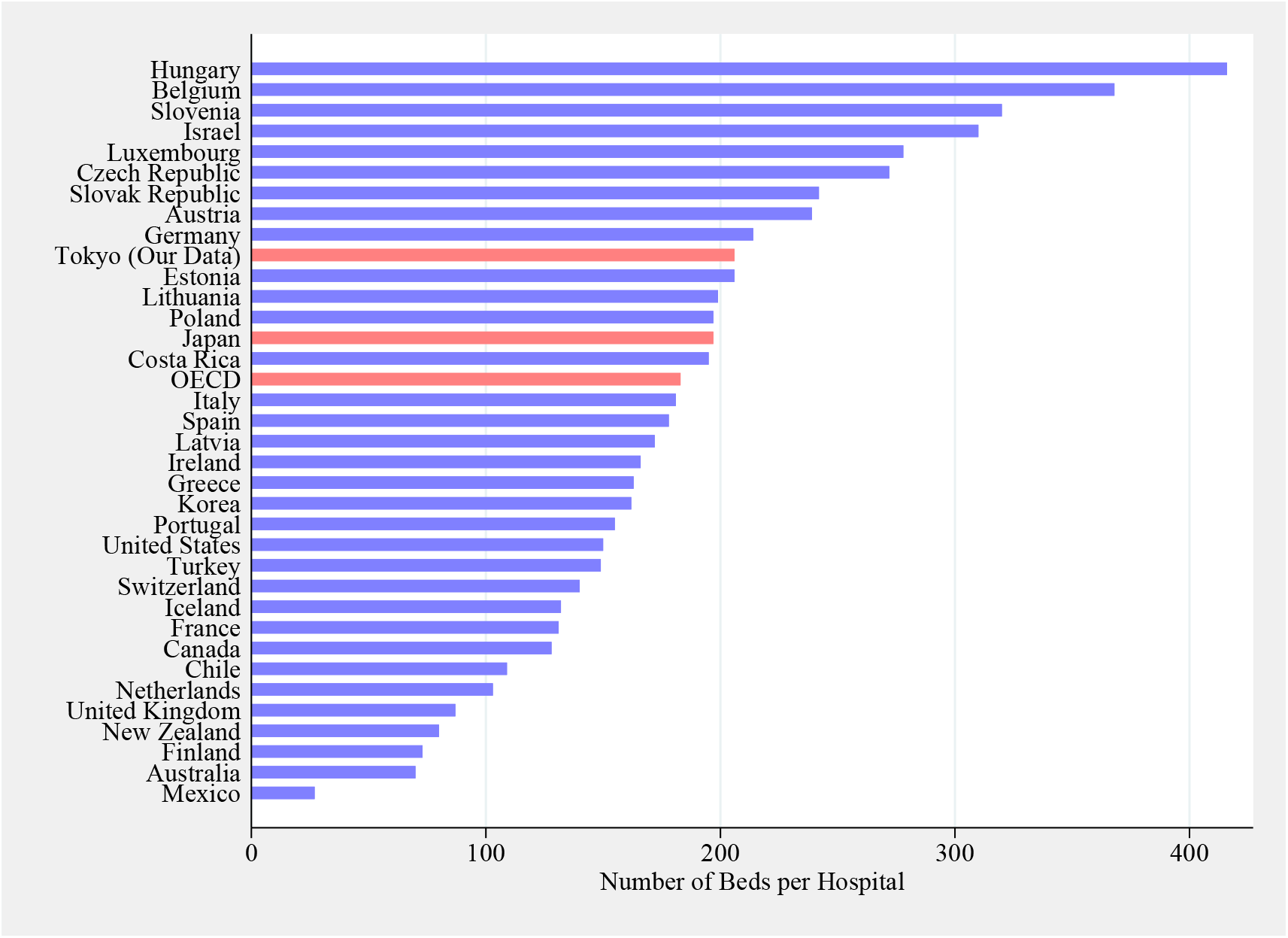
Hospital Size in OECD Countries *Notes:* The data are as of 2017. The average of OECD countries is derived as the arithmetic mean of 34 countries appeared in this figure.

### A.2 The COVID-19 Outbreak in Tokyo

The first case of COVID-19 in Tokyo was confirmed in a Chinese man on January 24, 2020. The first case of infection in Tokyo residents who had not been to China was confirmed on February 13. Since then, the number of COVID-19 patients has rapidly increased. Table A is a timeline of the major events related to the outbreak of COVID-19 from January to May 2020.

**Table A:** COVID-19 Timeline of Major Events in 2020 *Notes:* PG refers to the Prefectural Governor Yuriko Koike. Data and events were obtained from various sources.

| Date | Events |
| --- | --- |
| 24-Jan | The first COVID-19 case was confirmed in Tokyo. |
| 29-Jan | Charter flights from Wuhan arrived at Haneda airport. |
| 18-Feb | The Tokyo local government adopted the first supplementary budget. |
| 19-Feb | Passengers of the Diamond Princess ship who had tested negative began to disembark. |
| 20-Feb | The prime minister requested all schools to be closed from March 2. |
| 16-Mar | 140 hospital beds were secured in Tokyo. |
| 24-Mar | Tokyo Olympics was postponed. |
| 25-Mar | PG requested people to stay indoors on weekends. |
| 30-Mar | PG requested self-restraint in visiting karaoke shops, bars, and nightclubs. |
| 1-Apr | 620 hospital beds were secured in Tokyo. |
| 2-Apr | 700 hospital beds were secured in Tokyo. |
| 3-Apr | PG requested hospitals to provide beds for COVID-19 patients (Total 1700 beds). |
| 5-Apr | PG announced that 1,000 hospital beds were secured in Tokyo. |
| 7-Apr | The prime minister announced a state of emergency in 7 prefectures.<br>PG requested people to strictly observe stay-at-home orders.<br>One hotel in Tokyo was converted into an accommodation facility for COVID-19 patients. |
| 9-Apr | PG requested hospitals to provide beds for COVID-19 patients (Total 3300 beds). |
| 15-Apr | PG announced that 3,000 COVID-19 patients with mild symptoms could stay at accommodation facilities. |
| 16-Apr | Nation-wide state of emergency began. |
| 21-Apr | 2,000 hospital beds were secured in Tokyo. |
| 23-Apr | PG requested people to stay at home over long consecutive holidays from April 25 to May 6. |
| 5-May | Nation-wide state of emergency was prolonged to May 31.<br>400 hospital beds were secured for severely ill patients, 1600 beds for patients with moderate symptoms. |
| 14-May | State of emergency was lifted in 39 prefectures. |
| 25-May | State of emergency was lifted in Tokyo and other prefectures. |

To provide adequate care for COVID-19 patients and prevent hospital staff and non-COVID-19 patients from contracting the infection, the Ministry of Health, Labour and Welfare (MHLW) first ordered COVID-19 patients to be admitted to a Type 2 Designated Medical Institution for Infectious Diseases (hereafter, Type2-DMIID hospitals) in early February.^17^The number of beds available for COVID-19 patients in Type2-DMIID hospitals, however, was only 522 (MHLW, 2019), which was far from sufficient.

Therefore, on February 9, 2020, the MHLW released guidelines regarding the requirements for general hospitals (i.e., hospitals other than Type2-DMIID hospitals) for admitting COVID-19 patients. In these brief guidelines, the MHLW first stated that COVID-19 patients should be admitted to Type2-DMIID hospitals as often as possible. Next, the MHLW listed the three requirements (described in the main text) for general hospitals admitting COVID-19 patients. For non-Japanese readers, a full English translation of these guidelines is provided below.

As explained in the guidelines, details about the third requirement are available from the “Standards for Medical Institutions as Designated by the Minister for Health, Labour, and Welfare based on Article 32, Subsection 2 of the Act on the Prevention of Infectious Diseases and Medical Care for Patients with Infectious Diseases” as well as the “Guidance on Facility Standards for Medical Institutions Designated for Infectious Diseases.”

#### Securing beds for patients infected with the novel coronavirus (Request)

Regarding the novel coronavirus (referring only to the coronavirus, the pathogen of which is the beta coronavirus, limited exclusively to the disease that was newly reported to the World Health Organization by the People’s Republic of China in January 2020 and which has the ability to be transmitted to humans), through the enactment of the Government Ordinance Stipulating the Novel Coronavirus as a Designated Infectious Disease (Ordinance 11, 2020, the “Designation Ordinance,” the Government Ordinance to Amend the Quarantine Law Enforcement Ordinance (Ordinance 12, 2020), and the provisions of Article 3 of the Government Ordinance Stipulating the Novel Coronavirus as a Designated Infectious Disease, the Ministerial Ordinance on the Replacement of Terms when Applying the Provisions of Law Enforcement Regulations on the Prevention of Infectious Diseases and Medical Care for Patients with Infectious Diseases (MHLW, Ordinance No. 9, 2020), as well as the Ministerial Ordinance to Partially Amend the Quarantine Law Enforcement Regulations (MHLW, Ordinance No. 10, 2020), have now been enforced.

A cruise ship recently docked at a port in Yokohama City in Kanagawa Prefecture. Based on the fact that a large number of patients infected with the coronavirus were reported on this cruise ship, we have compiled the following list of medical institutions to which patients suffering from the coronavirus were sent. Therefore, once you have familiarized yourself with it, please inform relevant parties.

Regarding this request, based on the fact that there are a great number of reports of people being infected with coronavirus in different areas at this time, this request is a tentative one and we wish to add that it should not be considered as impacting infectious disease prevention policy in ordinary times.

1. Based on the Designation Ordinance and the Act on the Prevention of Infectious Diseases and Medical Care for Patients with Infectious Diseases (Law no. 114., 1998, the “law,” regarding patients infected with the coronavirus and those with related symptoms, in Principle, they must be hospitalized and placed in an infectious disease bed at a designated medical institution for infectious diseases. However, in the proviso to Article 19 Subsection 1 of the law, in emergencies or in other unavoidable circumstances, it is possible that a patient is hospitalized and placed in a bed other than one designated for infectious diseases, or in medical institutions other than those designated for infectious diseases.
2. Specifically, in case of transporting patients infected with the coronavirus to medical institutions, please be aware of the following points:

- Even in the event that circumstances correspond to the proviso of Article 19 Subsection 1 of the law, the patient will be transported to a designated medical institution for infectious diseases (However, there is no need to place them in an infectious disease bed.).
- With due cognizance that medical institutions must provide treatment for infectious diseases to residents in each region, the following points must be adhered to in the event that patients infected with the novel coronavirus are hospitalized in facilities other than designated medical institutions for infectious diseases, or in the event that they are placed in beds other than infectious disease beds at designated medical institutions for infectious diseases:

1). Ideally patients would be placed in private rooms when hospitalized. However, for groups of patients with a confirmed diagnosis of the novel coronavirus, they may all be treated in the same room.
2). Leveraging facilities like portable toilets, inpatients infected with coronavirus should not share toilets with other patients.
3). In addition, the procurement of appropriate beds with reference to the “Standards for Medical Institutions as Designated by the Minister for Health, Labour, and Welfare based on Article 32, Subsection 2 of the Act on the Prevention of Infectious Diseases and Medical Care for Patients with Infectious Diseases” (MHLW Notification No. 43, March 19th, 1999) and also the “Guidance on Facility Standards for Medical Institutions Designated for Infectious Diseases” (Notification from the Director of the Tuberculosis and Infectious Disease Control Division, Health Service Bureau, MHLW to the Directors of each Prefectural Health Department (Bureau).

Early in the 2020 outbreak, on February 20, Prime Minister Shinzo Abe requested all schools to be closed as of March 2, hoping for immediate containment of the infection. The Japanese government and an expert committee also recommended wearing face masks and avoiding the three Cs (closed spaces with poor ventilation, crowded places, and close-contact settings) right at the outset. Probably because of these proactive measures and people’s high compliance rates, the infection rate in early March was successfully suppressed. While Japanese media at that time argued that the low infection rate in Japan was simply due to the low rate of testing, the number of serious cases remained at a low level in early April, as shown in Figure A2.

**Figure A2:**
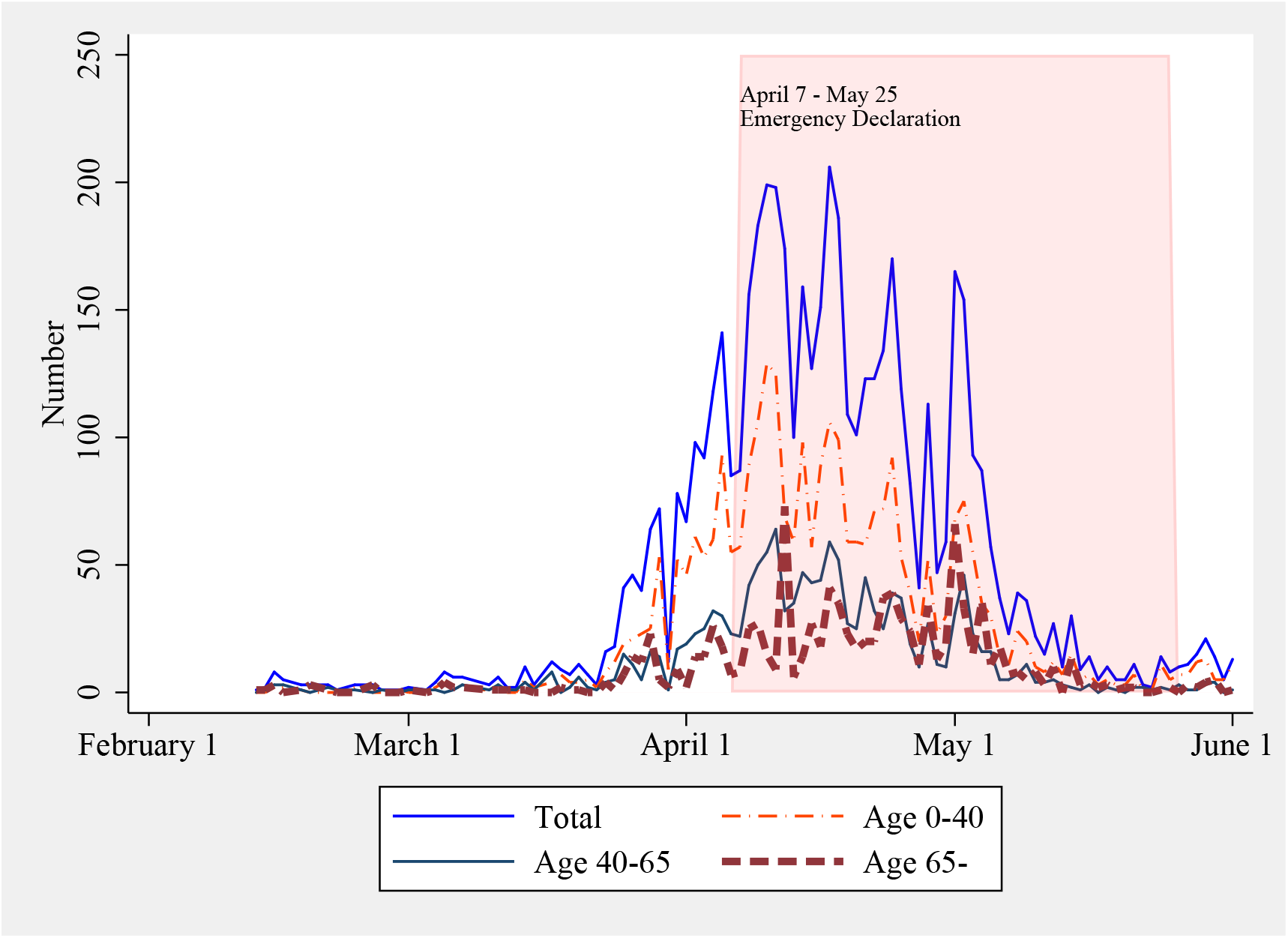
Number of Confirmed Cases of COVID-19 in Tokyo *Notes:* The colored area indicates the period during which a state of emergency was invoked.

Furthermore, there is wide regional variation in infection rates. Figure A3 shows a map of Tokyo shaded according to the infection rate in April 2020, when the number of new confirmed cases was at its peak. In general, the infection rate is high in the central area of Tokyo. Hospitals are mostly located in high-infection areas, where the population density is also high.

**Figure A3:**
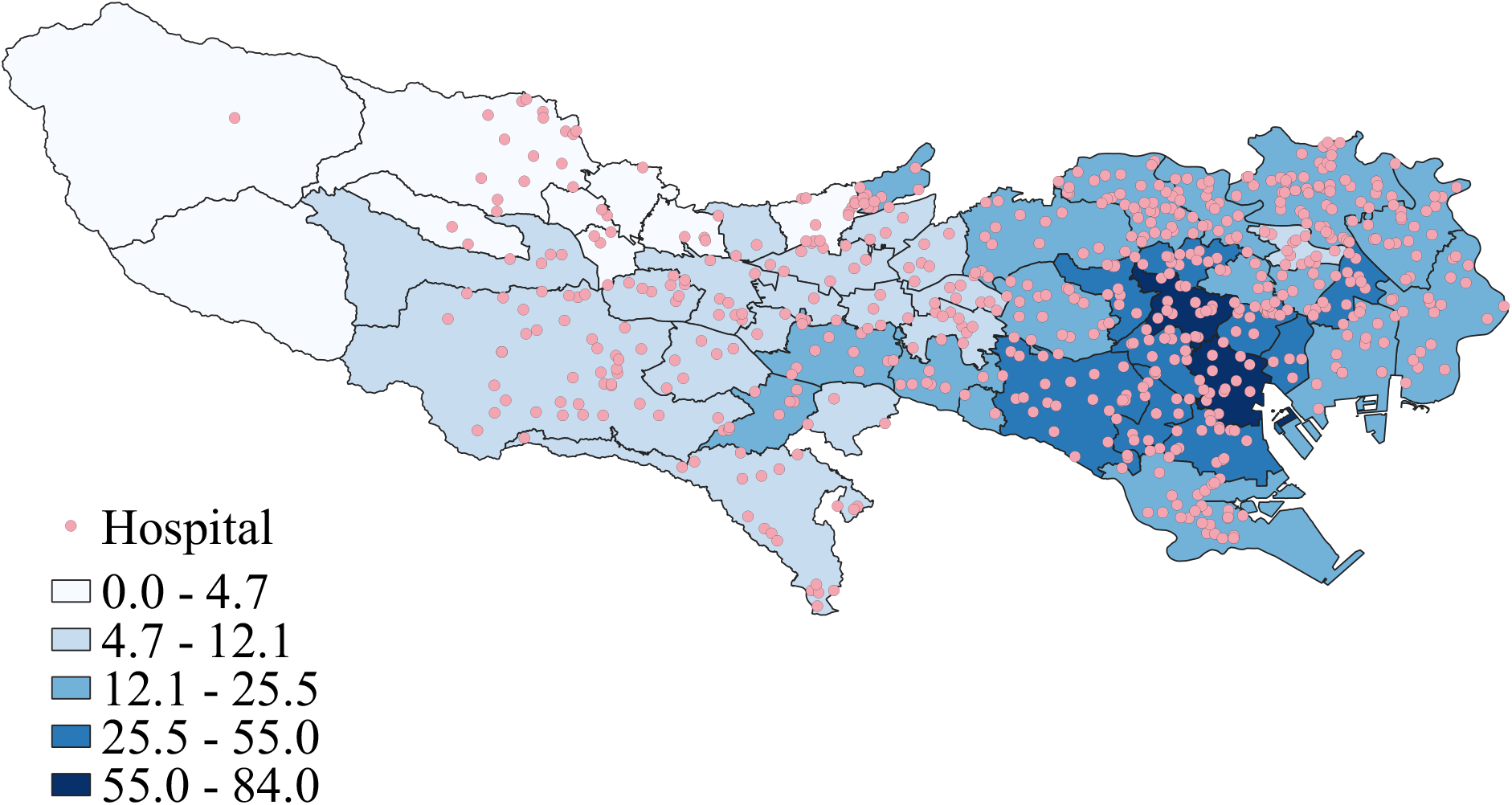
City-Level Infection Rate and Location of Hospitals: April *Notes:* Infection rate is per 100,000 people and based on the number of new confirmed cases in April. Each circle represents the location of a hospital.

Figure A4 shows how the Tokyo prefecture secured beds for COVID-19 patients. Although the Tokyo Metropolitan Government had secured only 140 beds by March 16, including 118 beds in Type2-IDDH hospitals, the number of secured beds increased rapidly in early April following the spread of infection. On March 26, the MHLW also ordered hospitals to postpone non-urgent elective procedures (MHLW, 2020a).

**Figure A4:**
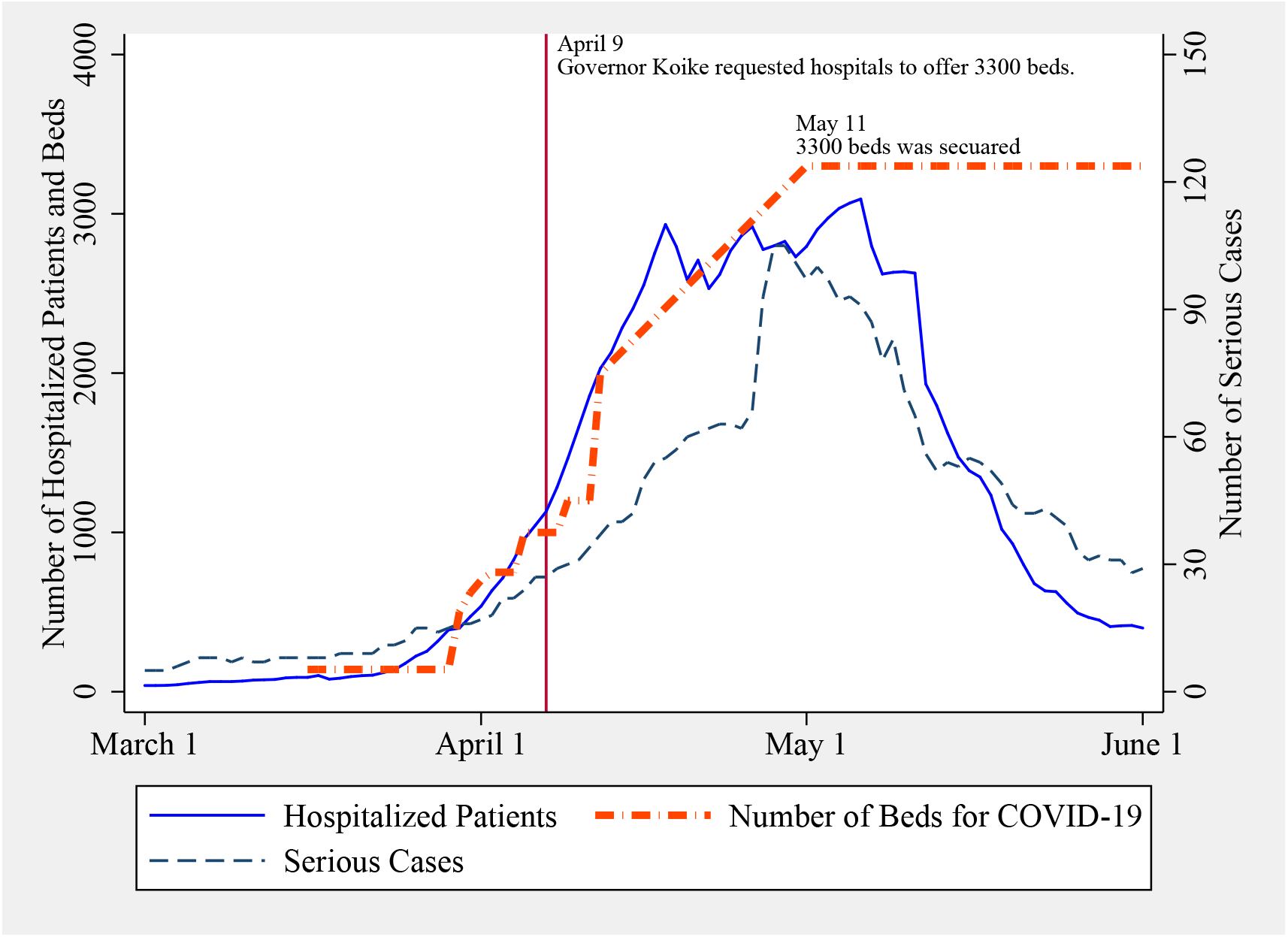
Number of inpatients and Secured Beds for COVID-19 Patients *Notes:* The secondary Y-axis represents the number of serious cases. The vertical line represents April 9th, when the Prefectural Governor Yuriko Koike requested hospitals in Tokyo to offer a total of 3,300 beds for COVID-19 patients.

To stop the rapid spread of infection, on March 25 Prefectural Governor Koike requested people to stay at home on weekends. Likewise, she requested self-restraint when visiting karaoke shops, bars, and nightclubs. Despite these measures, the number of confirmed cases increased and a state of emergency was declared in Tokyo starting April 7. On April 9, to handle the surge in COVID-19 cases, Prefectural Governor Koike again requested all hospitals in Tokyo to provide beds until the total number of beds for COVID-19 patients reached 3,300. In response to this request, 2,000 beds were secured by April 21.

In Japan, people generally travel during the long holidays from April 25 to May 5, the Golden Week. Because traveling during the Golden Week would have further spread COVID-19, on April 23 the prefectural governor strongly requested that people stay at home. On May 5, the state of emergency was prolonged until the end of May. Eventually, the state of emergency was lifted in Tokyo on May 25.

## B Trend of Main Outcome Variables

We checked the trend of the main outcomes without adjustment, in a manner similar to the standard checks for the common trend assumption in difference-in-differences /event study analysis. Figure B compares the trend in outcomes from February to May in both 2019 and 2020. Figure B(a) shows the results for the monthly profits per hospital bed. In this figure, square makers represent the data of hospitals that admitted COVID-19 patients (treatment group). Circle markers represent data from other hospitals (control group). Solid and dashed lines represent the data for 2020 and 2019, respectively.

**Figure B:**
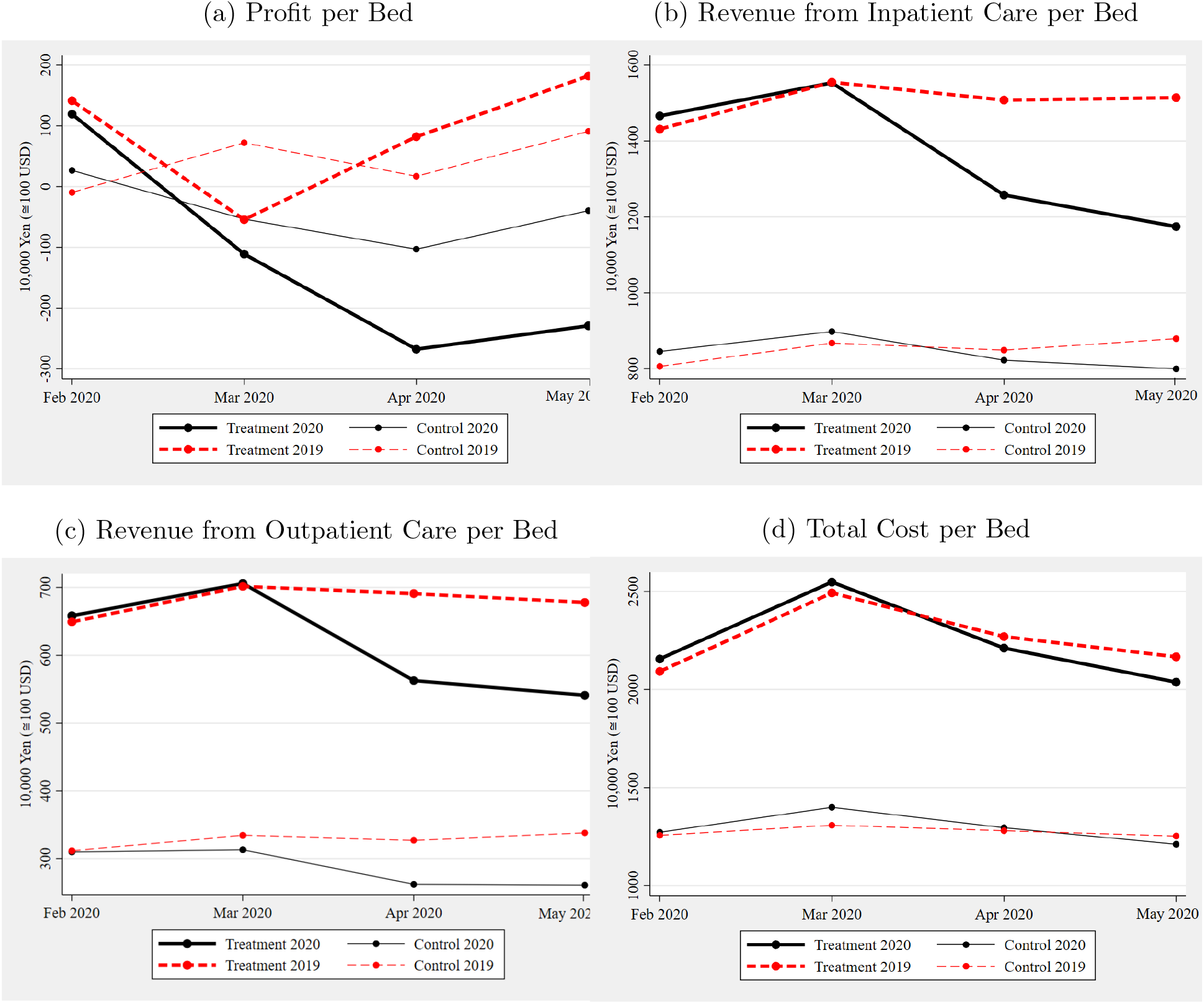
Trend in Selected Outcome Variables *Notes:* The treatment group consisted of hospitals that admitted COVID-19 patients from February to May 2020. The control group consisted of hospitals that did not. Solid and dashed lines represent the data for 2020 and 2019, respectively.

Note that the profits per bed in February 2019 and 2020 were almost the same in both groups. This suggests that the financial situation in February 2020, when COVID-19 had not yet spread in Japan, was similar to that in the previous year, regardless of whether the hospitals admitted COVID-19 patients. In hospitals with COVID-19 patients, however, we see a clear difference in the profit per bed between April/May 2019 and April/May 2020, with the 2020 profit JPY 300,000 lower. Furthermore, even in hospitals that did not admit COVID-19 patients, there was a substantial decline in profits in those months in 2020 because many patients refrained from visiting hospitals for fear of infection. By subtracting the difference in profits between April and May in the control group from that in the treatment group, we can roughly understand the effect of COVID-19 admissions. From this figure, we see that the treatment groups experienced an additional reduction in profits by JPY 150,000 to 200,000 in April–May. This amount is consistent with the OLS analysis in Table 3 on the effect of admitting COVID-19 patients.

The interpretation of the other outcomes follows a similar approach. In Figure B(b) and (c), the trends of revenue from inpatient care and outpatient care per bed are compared for the treatment and control groups in both 2019 and 2020. Clearly, the trends from February to March were similar in both the treatment and control groups, suggesting that the trends would have been sufficiently parallel from April to May in the absence of the COVID-19 outbreak. In the treatment group, however, we saw sharp reductions in revenues in 2020.

In Figure B(d), the trends in total costs are compared. While the decline in medical revenue also results in a similar decline in costs, we only find an ambiguous reduction in total costs in this figure. That is because personnel costs, which account for about 60% of total hospital costs in normal times, could not be adjusted immediately even if medical demand for hospital care exhibited a sharp reduction.

## C Derivation of Complier Characteristics

### C.1 Conceptual Framework

Before we explain the detailed procedures for deriving complier characteristics, we discuss Heckman’s conceptual framework (Heckman et al, 1997; Heckman and Vytlacil, 2001), which has been recently applied to health insurance policies (Kowalski, 2016; Abrigo et al, 2019). We let *D* represent a binary treatment of whether to admit COVID-19 patients and let *Y* represent an observed outcome, such as profits per bed. *Y_T_* and *Y_U_* are potential outcomes with and without admitting COVID-19 patients. With panel data, we use different specifications. We consider two time periods, where *Delta* represents changes between “before” and “after.” With these notations, a change in the observed outcome as a result of the COVID-19 outbreak (Δ*Y*) is related to potential outcomes using the following equation:

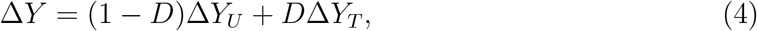

where Δ*Y_U_* and Δ*Y_T_* are potential changes when hospitals admit or do not admit COVID-19 patients.

Notably, there is a substantial selection of the treatment (*D*) in the framework. For example, there is substantial heterogeneity in the efficiency of hospitals dealing with the surge in COVID-19 patients. Some hospitals were willing to admit COVID-19 patients with mild symptoms because they could transfer them to other hospitals when they started exhibiting serious symptoms, thus minimizing financial damage. In contrast, many hospitals reluctantly admitted COVID-19 patients out of fear of suffering catastrophic financial damage otherwise.

Therefore, we identify the effect of admitting COVID-19 patients using an instrumental variable estimation, while explicitly considering that the status of whether to admit COVID-19 patients is a result of an endogenous decision. Policymakers are also interested in the effect of admission on compliers (Imbens and Angrist, 1994).

In this model, a hospital selects treatment *D* if the net benefit of admitting COVID-19 patients (*I_D_*) is greater than or equal to zero. *I_D_* consists of an observed component *p* and an unobserved component *U_D_* as follows:

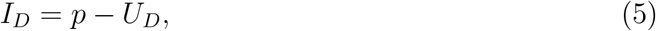

where *p* represents the benefits of admitting COVID-19 patients, which are influenced by instrumental variables (*Z*), and *U_D_* represents the unobservable costs, which are assumed to be uniform random variables. Admitting COVID-19 patients involves the benefit of avoiding an incessant bad reputation. In fact, hospitals in Japan are strongly motivated to avoid a bad reputation, which would have been inevitable if they had completely turned away COVID-19 patients and continued to provide only the usual medical care. Since the Tokyo Metropolitan Government has great authority to control the number of hospital beds in Tokyo, hospitals with a bad reputation would have found it difficult to increase capacity in the future. Thus, while admitting COVID-19 patients results in a financial loss, avoidance of a bad reputation is a significant benefit.

As *p* differs across hospitals, hospitals are assumed to have different values of *U_D_*. For example, hospitals with altruistic motivations to contribute to the care of COVID-19 patients, motives that cannot be observed, may have low *U_D_*.

In the simplest case with a binary instrument, *p* takes one of two values (*p*_0_ and *p*_1_) for hospitals with *Z* = 0 and *Z* = 1, respectively. We can also calculate the proportion of compliers as (*p*_1_ *− p*_0_) *∗* 100%,^18^ and the local average treatment effects (LATE) on a detrended outcome are defined by the following equation:

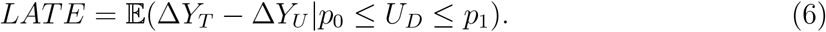

In this framework, hospitals with *U_D_* larger than *p*_0_ but lower than *p*_1_ always receive treatment if *Z* = 1 because their benefit from admitting COVID-19 patients exceeds costs. By contrast, if *Z* = 0, they never admit COVID-19 patients. In other words, the decision to admit COVID-19 patients is always consistent with *Z* when *U_D_* is within the interval of *p*_0_ to *p*_1_. The LATE uncovers the average treatment effects within this particular.

### C.2 Derivation of Complier Characteristics

As mentioned previously, our IV estimator identified the effect of COVID-19 admissions for a specific hospital group—the compliers—whose treatment status (*D*) is determined by instruments (*z*). To properly interpret the LATE and understand the mechanisms, it is important to consider the characteristics of compliers.

To do so, we followed the methodology proposed by Kowalski (2016). We first synthesized multiple continuous variables used in the main analysis into a binary variable by extracting the first component from the results of a principal component analysis. Based on the median of the first component, a binary instrumental variable (*z*) was constructed. While this transformation is somewhat it is helpful in capturing the variation caused by instrumental variables.

This binary instrumental variable allows us to discuss how the instrumental variable is related to the treatment status and to show it graphically. In figure C, always-takers (“unsuitable” hospitals that admit COVID-19 patients) are those who do not receive intervention (*Z* = 0) but still provide treatment. Assuming a uniform distribution of *U_D_*, the fraction of always-takers is given as *p*_0_ = 𝔼(*D* = 1*|Z* = 0). Note that we can never identify always-takers individually. For example, some of those with *D* = 1 and *Z* = 1 may also be always-takers because they would have admitted COVID-19 patients even if they were unsuitable hospitals. In contrast, hospitals with *D* = 1 and *Z* = 0 can be correctly identified as always-takers. Therefore, by using the characteristics of hospitals with *D* = 1 and *Z* = 0, it is possible to infer the average characteristics of always-takers.

**Figure C:**
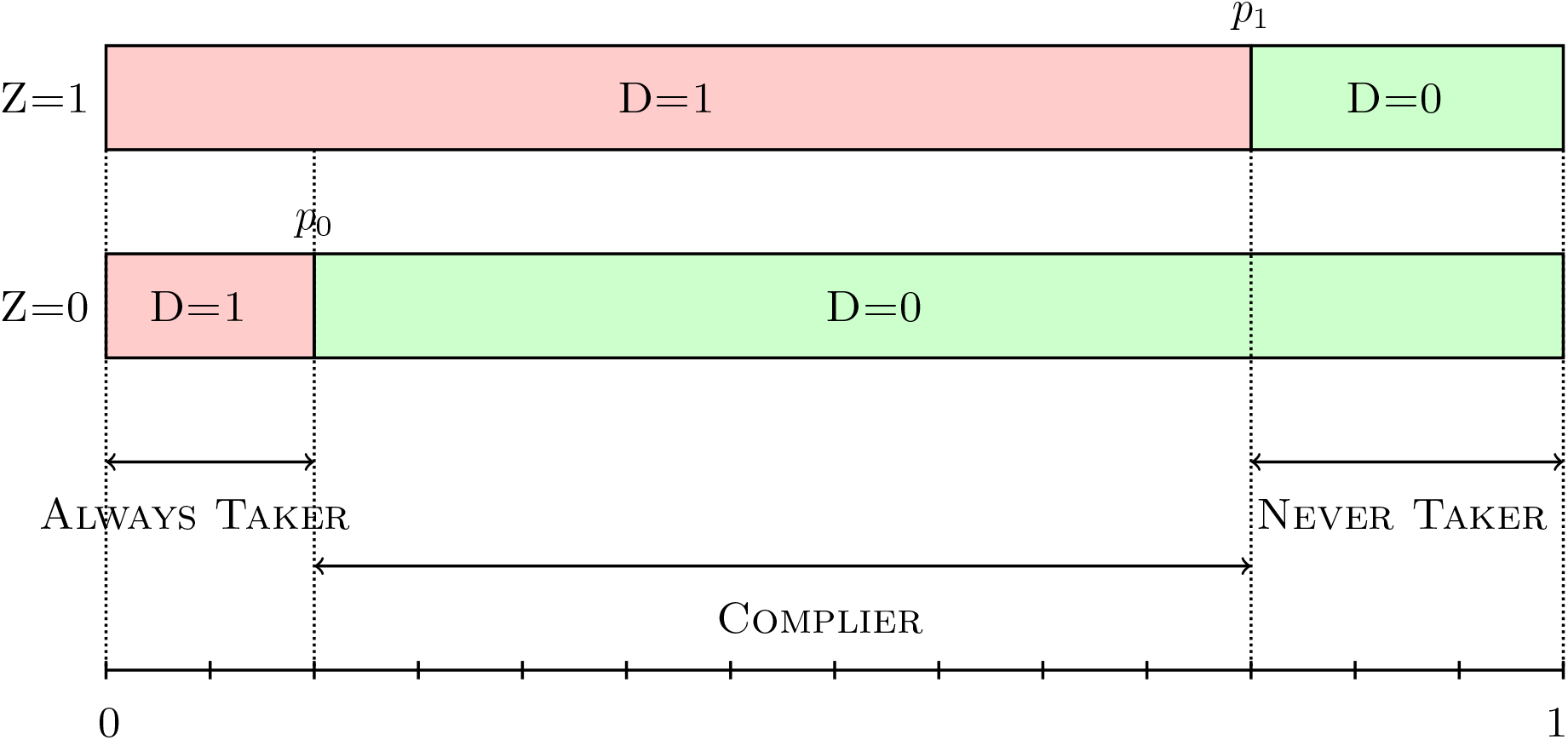
Identification of the Complier *Notes:* The treatment *D* is a binary variable, which is equal to 1 if a hospital admitted COVID-19 patients during the study period, and 0 otherwise. The instrument *Z* is equal to 1 if the first principal component, which measures how the hospital could suitably admit COVID-19 patients, exceeded the median. *p*_0_ is determined as the fraction treated in unsuitable hospitals (*P*(*D* = 1| = 0)), and *p_1_* is determined as the fraction treated in suitable hospitals (*P*(*D* = 1|*Z* = 1))

The parallel discussion holds for never-takers who did not admit COVID-19 patients regardless of whether they were suitable hospitals. Here, hospitals with *D* = 0 and *Z* = 1 are correctly categorized as never-takers, while some hospitals with *Z* = 0 may also be never-takers. Assuming a uniform distribution of *U_D_*, the fraction of always-takers is given as *p*_1_ = 𝔼(*D* = 0*|Z* = 1).

Finally, we considered hospitals with *p*_0_ *≤ U_D_ ≤ p*_1_. Their decision to admit COVID-19 patients was completely consistent with the assignment of the instrumental variable. For example, if *Z* = 1, their propensity to offer treatment was *p*_1_; thus, they decided to be in the treatment group because their unobservable costs (*U_D_*) were less than the benefits (*p*_1_). By contrast, if *Z* = 0, they did not offer treatment because *U_D_ > p*_0_. In this sense, hospitals with *p*_0_ *≤ U_D_ ≤ p*_1_ were categorized as compliers.

In the actual implementation to determine the values of *p*_1_ and *p*_0_, we estimated the first-stage regression with this newly created instrument and obtained the predicted values. These predicted values give the intervals that define always-takers, compliers, and never-takers Kowalski (2016). In our article, *p*_1_ is set at 𝔼(*D|Z* = 1*, X*) and *p*_0_ is also set at 𝔼(*D|Z* = 0*, X*). After running the first-stage regression, we obtained 0.1268 for *p*_0_ and 0.4865 for *p*_1_, respectively. With these values and the assumption of a uniform distribution of *U_D_*, the fraction of compliers was estimated to be 35.97% (48.65% *−* 12.68%). The fraction of never-takers was 51.35% (100% *−* 48.65%).

After determining the values of *p*_1_ and *p*_0_, we estimated the average characteristics of compliers. First, we computed the average characteristics of always-takers and never-takers from 𝔼(*X|D* = 1*, Z* = 0) and 𝔼(*X|D* = 0*, Z* = 1).

Contrary to the average characteristics of always-takers and never-takers, the average characteristics of compliers cannot be directly observed. For example, we can observe 𝔼(*X|D* = 1*, Z* = 1), but this value reflects a mixture of the characteristics of always-takers and compliers because some hospitals with *D* = 1 and *Z* = 1 would have offered treatment even if they were not exposed to the binary instrumental variable (*Z* = 0). Therefore, the characteristics of treated compliers (*µ_X_* (1)) and untreated compliers (*µ_X_* (0)) are derived by subtracting the characteristics of always-takers (never-takers) from the composite characteristics of always-takers (never-takers) and compliers, as shown in equations 7 and 9 (Kowalski, 2016; Abrigo et al, 2019).

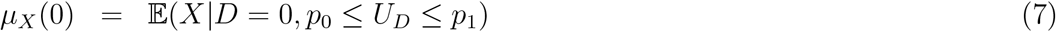

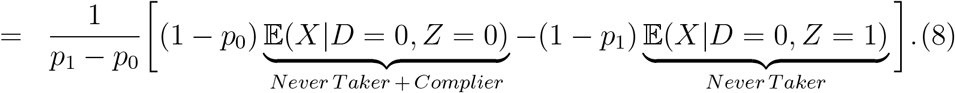

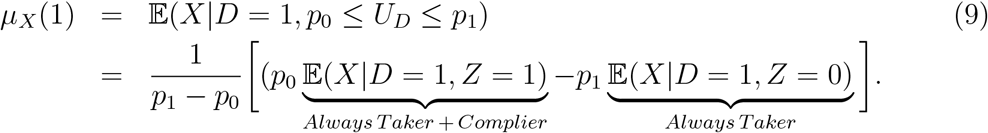

After calculating the average characteristics of the untreated and treated compliers separately, we obtained the characteristics of the compliers by calculating the weighted sum of these two players. Since we created a binary instrument based on the median, the weight was set to 50%. Finally, we replicated these procedures 1000 times with the bootstrap method and calculated the bootstrap standard errors as well as the point estimate for the mean.

## D Results on Other Outcomes

To supplement these results, we explored the effects on other outcomes. Table D summarizes the OLS and IV estimation results. The specification in Column (2) in Table 3 is used for the OLS estimations, while the specification in Column (6) in Table 3 is used for the IV estimations. Regardless of the regression method, the coefficient of *COVID* is significantly negative for all dependent variables, other than the number of outpatients. The trend in which the OLS estimate is larger (i.e., smaller negative) than the IV estimate can also be applied to Table D.

**Table D:**
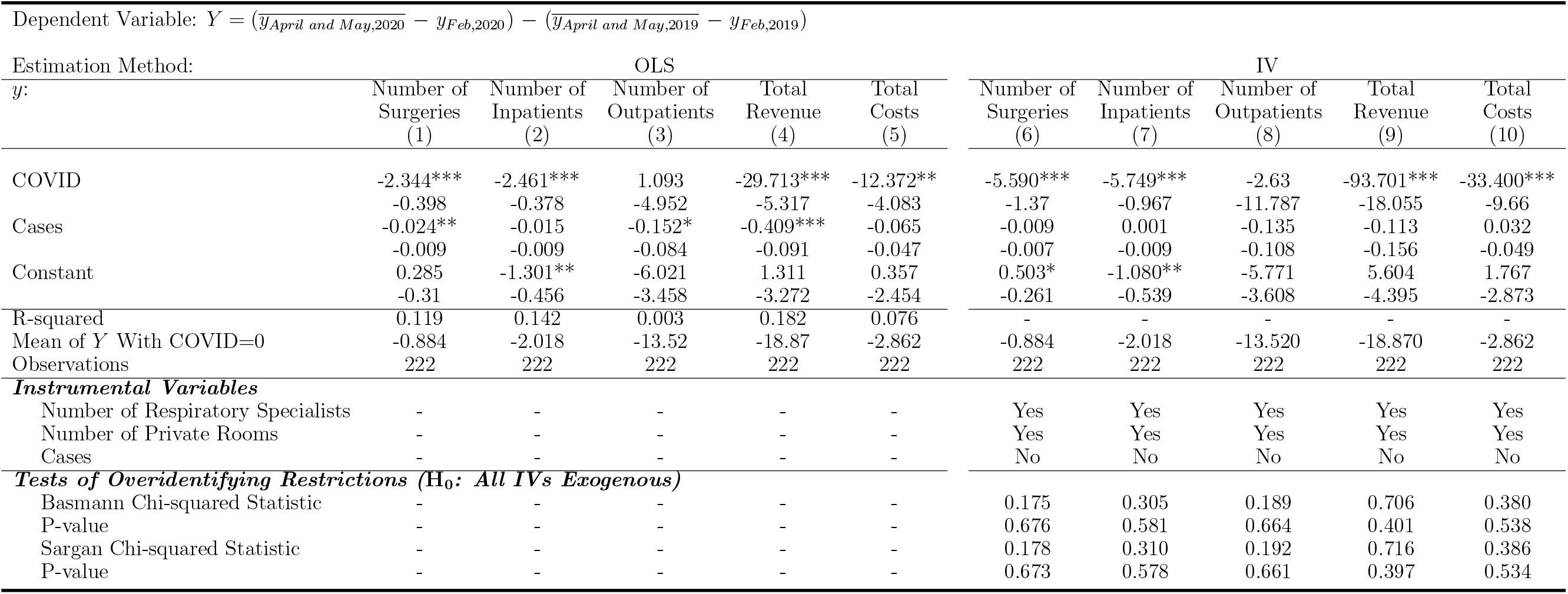
Results on Various Outcomes *Notes:* The dependent variable is the year-on-year differences in each variable, divided by the number of beds. The unit is 10,000 JPY. *Cases* is the monthly number of COVID-19 patients around each hospital. Standard errors clustered at the level of 12 medical areas are reported in parentheses. *** p<0.01; ** p<0.05; * p<0.1.

Again, according to the over-identifying restrictions tests, the results strongly suggest that all the IVs are exogenous for the IV estimations with these dependent variables as well. Thus, in terms of the exclusion restriction, the IV estimates were considered reliable.

We found a significant decrease in the number of surgeries performed per bed. The IV estimate for the coefficient of *COVID* was -5.590 and statistically significant. Since the average of this dependent variable for the control group (i.e., hospitals that did not admit COVID-19 patients) was -0.884, this treatment effect is remarkably large. While the MHLW requested postponing elective surgeries for all hospitals (MHLW, 2020a), hospitals that admitted COVID-19 patients postponed or canceled surgeries to a larger extent than did other hospitals.

The number of inpatients also decreased, and the estimated coefficient on *COVID* was statistically significant, even at the 1% significance level. While the hospitals without COVID-19 patients experienced a decrease of 9.224 inpatients per bed, the reduction in the number of inpatients in the hospitals with COVID-19 patients is much larger according to Table 1, which also implies a large reduction in medical revenue. Indeed, the estimated coefficient of *COVID* for the total revenue is -93.701, statistically significant even at the 1% significance level.

We also found a significant reduction in total costs. The decrease in medical activities necessarily reduced costs; however, personnel costs, which account for about 60% of total costs in many hospitals, were not quickly adjusted. Therefore, we found a larger decrease in total revenue than in total costs.

## E Robustness checks excluding hospitals with ICU

While our main results showed large negative effects on profits, those results may change when we consider the heterogeneous effects between hospitals with and without an intensive care unit (ICU).

In general, hospitals with ICUs provide very expensive care for COVID-19 patients with severe symptoms (Khan et al, 2020). Thus, we expected that the negative effect of admitting COVID-19 patients would be greater in these hospitals.

Figure E(A) plots the association between inpatient revenue per inpatient in 2019 and changes in profits (the main outcome in this study). In this figure, we find a clear negative association, suggesting that hospitals with higher revenue per patient in the previous year experienced a larger reduction in profits owing to the cancellation of their usual medical care. With some exceptions, the level of inpatient revenue per patient was similar between hospitals with and without an ICU. While hospitals with an ICU experienced greater financial damage from admitting COVID-19 patients, the extent of the differential effects between hospitals with and without an ICU may not be so large.

**Figure E:**
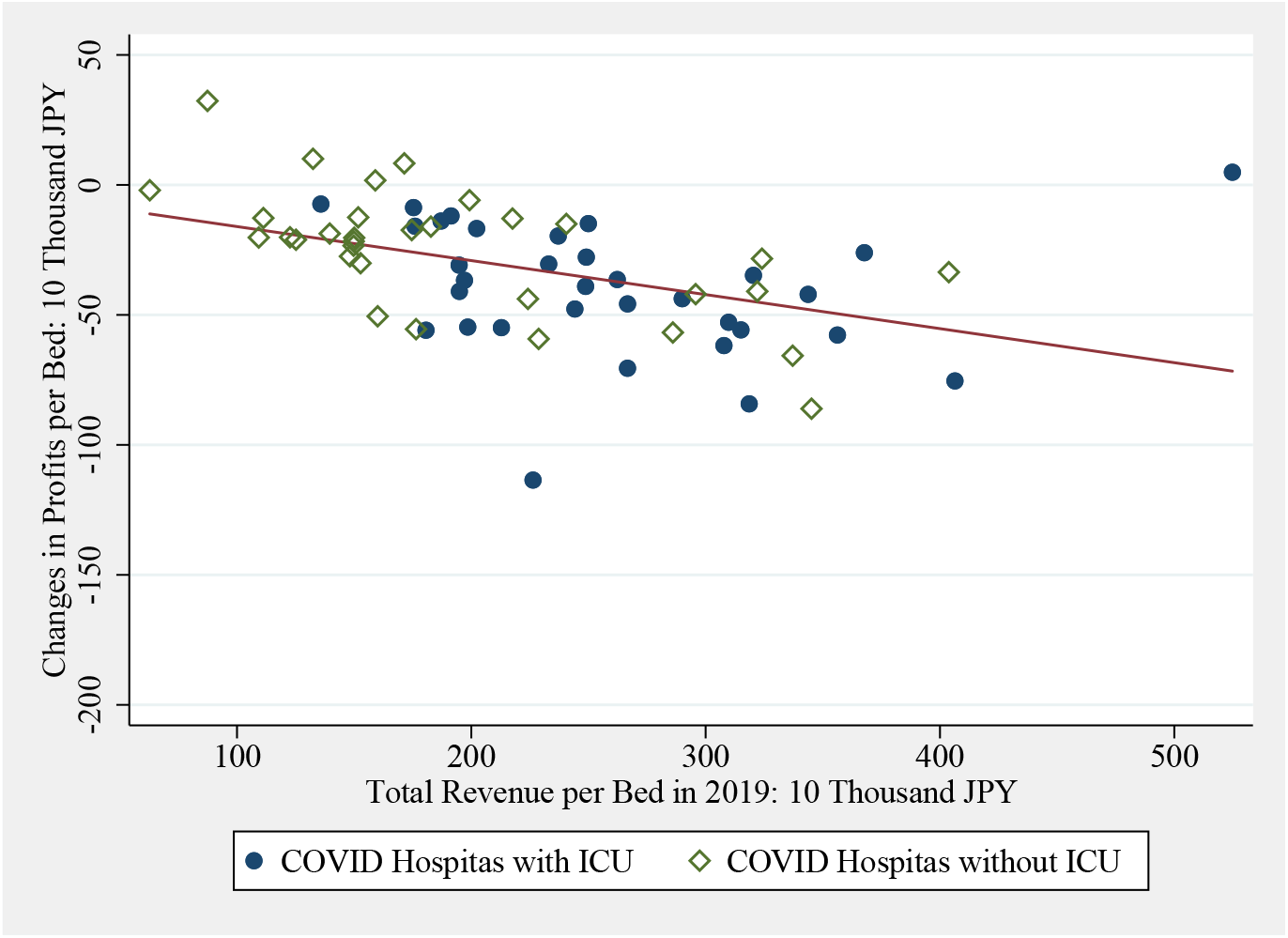
Changes in Profits among Hospitals with and without an ICU *Notes:* The sample consists of 68 hospitals that admitted one or more COVID-19 patients. The horizontal axis represents inpatient revenue per bed in 2019, and the vertical axis represents the changes in profits per bed from 2019 to 2020. The linearfit is shown as a solid line.

We also estimated the effect of admitting COVID-19 patients in hospitals without an ICU. The results are listed in Table E, which reports results from our preferred IV regression, using two IVs and controls for the local prevalence of COVID-19.

**Table E:** Results Excluding Hospitals with an ICU *Notes:* The dependent variable is detrended profits per bed. The unit is JPY 10,000. Standard errors clustered at the level of 12 medical areas are reported in parentheses. *** p<0.01; ** p<0.05; * p<0.1.

|  | (1) |
| --- | --- |
| COVID | -48.775*** |
|  | (16.828) |
| Cases | -0.1 |
|  | (0.145) |
| Constant | -3.761 |
|  | (4.095) |
| ***Instrumental Variables*** |  |
| Number of Respiratory Specialists | Yes |
| Number of Private Rooms | Yes |
| First-stage F Statistics | 6.803 |
| Mean of Dep. Var. among Hospitals with COVID=0 | -16.05 |
| Observations | 186 |

In Column (1), the cost effect of admitting COVID-19 patients was around JPY 487,000. This effect was slightly smaller than that from the entire sample, but the difference is not large. Therefore, the effect of admitting COVID-19 patients was still large even among hospitals that did not provide critical care in the ICU and admitted COVID-19 patients with mild or moderate symptoms.

While care for COVID-19 patients with severe symptoms is extremely costly, our results suggest that financial damage was also severe among hospitals that provided care for COVID-19 patients with mild symptoms. This suggests that the severe decline in profits was mainly driven by the cancellation of usual medical care, as confirmed in Online Appendix D, rather than by direct medical costs for COVID-19 patients.

## F Additional Placebo Test

As a useful intuitive check for unobserved confounders, we tested how the trend of our primary outcome (profits per bed) was associated with IVs during the pretreatment period. If our IVs had no direct effects on the outcome, we would find null effects when we regressed our IVs on the outcome before the COVID-19 outbreak.

The results are presented in Table F. In this table, the dependent variable is the year-on-year differences in profits per bed in each month. Columns (1) to (4) show the results from February through May. Note that the year-on-year differences in February work as a placebo test because in February there was no surge in COVID-19 patients.

**Table F:**
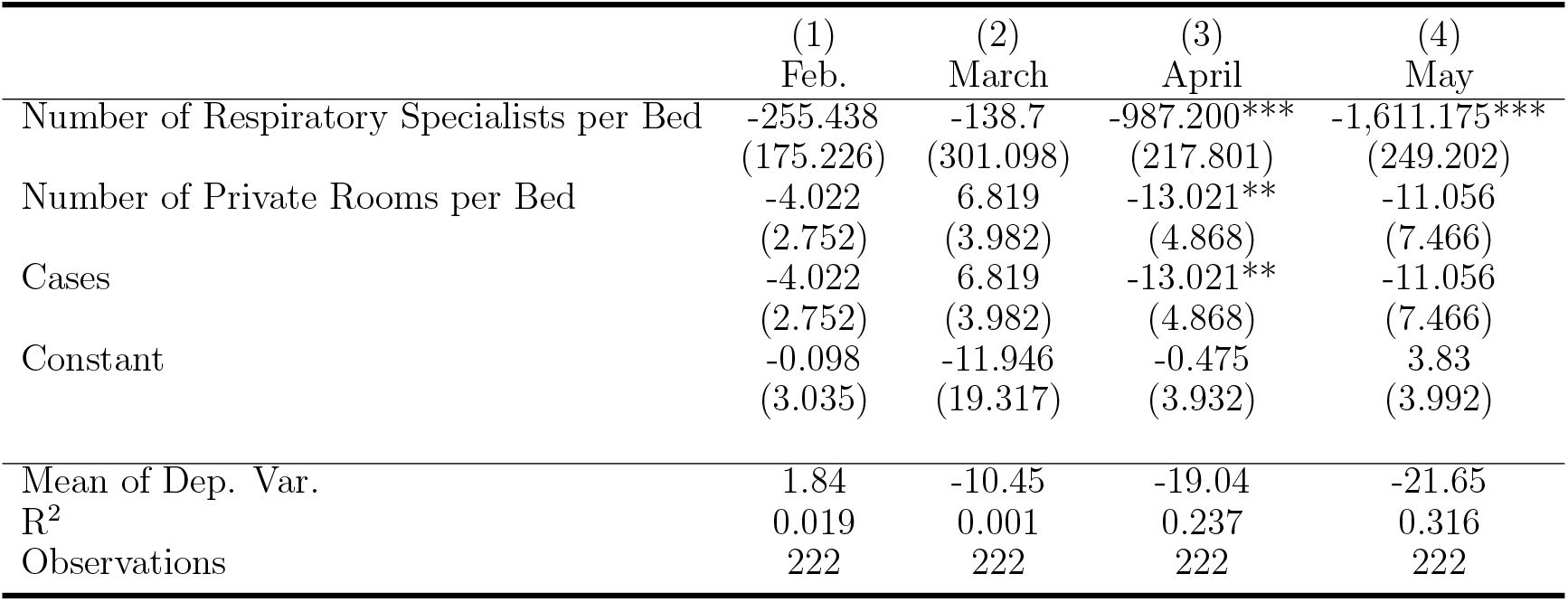
Additional Placebo Test *Notes:* The dependent variable is the year-on-year differences in profits per bed in each month. The unit is JPY 10,000. Standard errors clustered at the level of 12 medical areas are reported in parentheses. *** p<0.01; ** p<0.05; * p<0.1.

In Column (1), our IVs were not associated with the trend of profits per bed. The coefficients of the number of respiratory specialists and the number of private rooms were not statistically significant. This suggests that our IVs were not strongly associated with unobserved confounders. By contrast, the two IVs had strong predictive power in April and May when COVID-19 spread because they affected profits through the decision to admit COVID-19 patients.

## G Discussion of External Validity

We explored the external validity of our main results for unobservable characteristics, such as the priority placed on financial profit, because they determined whether hospitals admitted COVID-19 patients and may have had an effect on outcomes. For example, citetRosenthal2020 reported a typical example of “selfish hospitals.” According to her article, the phrase “No. 1 in COVID-19 Treatment” carries a stigma and, thus, is bad branding for some high-quality hospitals with a sufficient number of beds and protective equipment. For that reason, those hospitals did not admit COVID-19 patients. By contrast, “altruistic hospitals” admitted COVID-19 patients even at the risk of significant financial damage. Since these unobservable motivations to serve COVID-19 patients may also affect the influence on profits, we explicitly explored how the marginal effects varied by unobservable resistance to the treatment.

To do so, we estimated the marginal treatment effect (MTE) (Heckman and Vyt- lacil, 2005; Carneiro et al, 2015; Kamhöfer et al, 2019). According to Table 3, the use of any specification from Columns (3) to (6) is justifiable. We use Column (6) as the most preferable specification in the MTE analyses.

Figure G reports the common support and MTE curve for profits per bed. The exogenous variation from (i) the number of respiratory specialists per bed and (ii) the number of private rooms per bed creates common support in the propensity score over virtually the full unit interval. Note that it is very important to achieve nearly full common support for computing the ATT and the ATUT, which heavily weigh hospitals at the extremes of the treatment propensity distribution (Cornelissen et al, 2018). Note that ATT is the treatment effect calculated by more heavily weighting hospitals to the left of each Figure G, that is, hospitals with low unobserved resistance (*U_D_*) and a high propensity score (*p*), which results in *U_D_ < p*, making them accept COVID-19 patients. By contrast, ATUT is the treatment effect calculated by more heavily weighting hospitals to the right of each Figure G, that is, hospitals with high unobserved resistance (*U_D_*) and a low propensity score (*p*), which results in *U_D_ > p*, making them *not* accept COVID-19 patients.

**Figure G:**
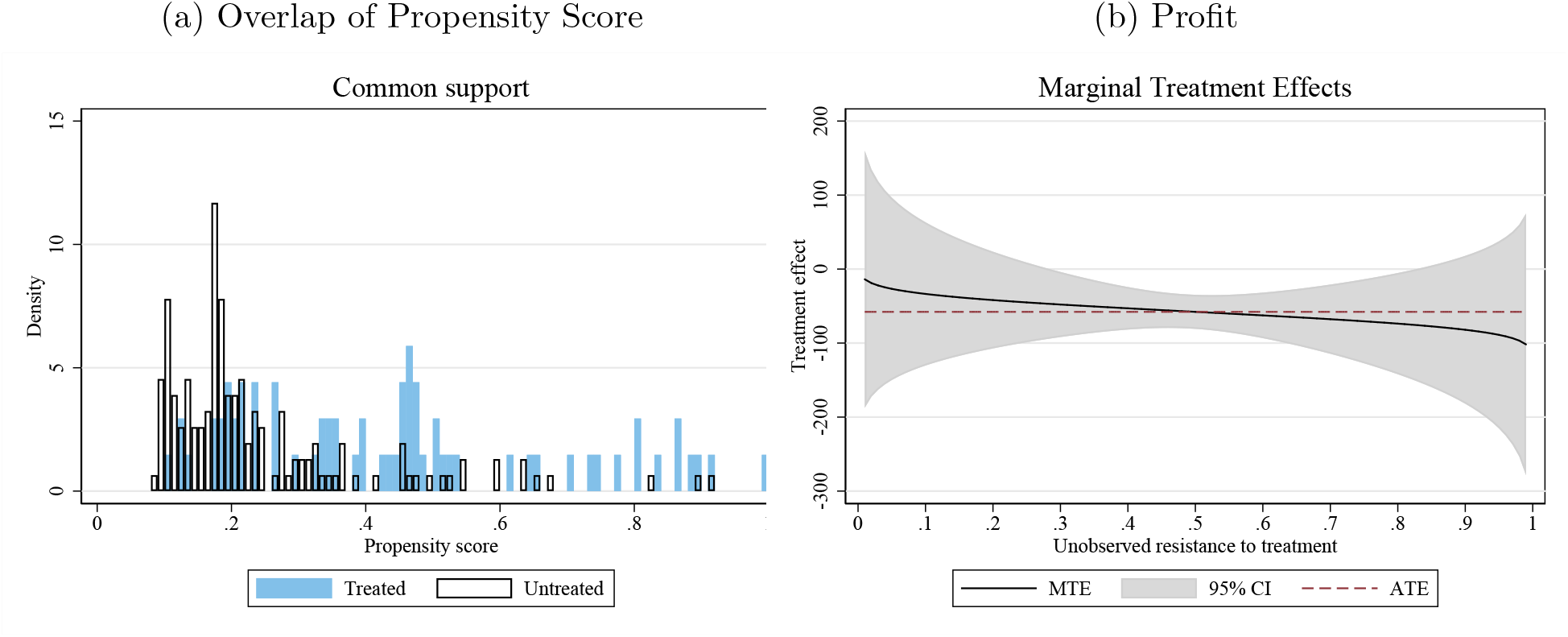
Common Supports and MTE Curve for Profits *Notes:* MTE curve is derived from the same specification as Column 6 in Table 3.

We noticed that the MTE curve of profits per bed was almost flat for the level of unobserved resistance to admitting COVID-19 patients. Furthermore, we could not reject the null hypothesis on the heterogeneity of treatment effects over unobserved resistance. This suggests that our results based on LATE have relatively high external validity for any level of unobserved heterogeneity in admitting COVID-19 patients.

The flat MTE curve in the profits makes sense because the preparations hospitals had to make to admit COVID-19 patients—such as the cancellation of surgeries and preparation of private rooms—would not differ according to the level of unobserved resistance, such as selfishness and altruism, even if the financial damage was highly heterogeneous for other characteristics of hospitals.

## Footnotes

1 From the perspective of classical health economics, sudden deterioration in hospital margins may have serious adverse effects on the quality of care. For example, hospitals become more likely to provide unnecessary and expensive care to compensate for the financial losses from admitting COVID-19 patients (Gruber and Owings, 1996; Yip, 1998).

2 Some reports do estimate hospital profit lost from COVID-19 admissions (American Hospital Association, 2020; Japan Hospital Association, 2020; GHC, 2020), but those estimates are the average effect among hospitals that admitted COVID-19 patients and do not consider hospitals’ heterogeneity.

3 Note that given this situation, we can divide all hospitals into three types; those who absolutely serve for COVID-19 patients as if there was no other option, the opposite type, that is, hospitals that never admit COVID-19 patients, and the remaining middle type who may or may not admit COVID-19 patients. For simplicity, let us call the last type of hospitals “swing hospitals” to resemble the phrase “swing states” in US elections, in the sense that both options could potentially be realized.

4 In terms of econometrics, swing hospitals are equivalent to compliers. These hospitals are “swinging” between two options, unlike hospitals that never admit COVID-19 patients (“never-takers”) or hospitals that always admit them (“always-takers”).

5 The most notable example is in Wuhan, China. The government of China rapidly built a 1,000- bed hospital, Wuhan Huoshenshan Hospital, in under 10 days, and immediately began caring for COVID-19 patients (Zhu et al, 2020). Following this approach, many countries such as the UK, Italy, and the US have also established temporary hospitals (Sacchetto et al, 2020).

6 Note that many researchers and spokespersons from medical organizations in Japan, such as (TMA, 2020) and (Kobayashi, 2020), also insisted on treating most COVID-19 patients in large public hospitals.

7 More details and an English translation of this guideline are shown in Online Appendix A

8 These numbers are tentative data provided by the Tokyo Metropolitan Government.

9 Even the Tokyo Medical and Dental University Medical Hospital, which admitted the largest number of COVID-19 patients, offered only 56 beds for COVID-19 patients out of 753 beds in total.

10 Since we use the double differences of *Y* as an outcome, the estimation of Equation 1 is essentially the same in the triple difference analysis that uses the level of *Y* as an outcome. Online Appendix B shows that the trend of the outcome variables seems parallel. *α*_1_ in Equation 1, however, measures the effect of admitting COVID-19 patients among hospitals that did so; thus, it is different from the effect we were interested in, as will be explained later.

11 *Case* is calculated with inverse 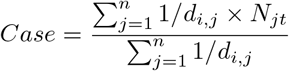 where *d_i,j_* is the distance from hospital *i* to city *j*, and *N_jt_* is the number of COVID-19 patients per 100,000 people in city *j* in April and May.

12 Note that the local prevalence of COVID-19, “*Case*,” can affect the potential number of patients when the hospital admits COVID-19 patients and hence the decision to admit such patients as well.

13 Nihon Ultmarc gathers detailed information on clinically active physicians for medical representatives (MRs), whose job is to meet with physicians and provide them information on pharmaceutical products. When MRs meet physicians, they report the physicians’ basic information to Nihon Ultmarc, who then immediately shares the information with all MRs registered in their system. In this way, Nihon Ultmarc successfully gathers information for almost all clinically active physicians in Japan.

14 The direct method to estimate the average characteristics of compliers for multiple continuous IVs has not been proposed yet, but it is still useful to discuss the average characteristics of compliers by transforming multiple IVs into one binary IV.

15 According to Khan et al (2020), on average, the cost per COVID-19 patient in the intensive care unit (ICU) is about twice that per patient in the general medical ward.

16 Note that what we mean by intervention here includes not only the governments’ request but also all the policy-related manners including the system that allows hospitals to decide whether to accept COVID-19 patients by themselves and the contents of the guideline issued by the MHLW.

17 Japan categorizes the severity of infectious diseases into five groups according to the risk each disease poses to public health. Type 1 refers to a very serious infectious disease for which no effective medical treatment has been established, such as the Ebola virus. Type 2 is for less severe diseases than Type 1 that still seriously impact public health, such as COVID-19.

18 Note that *U_D_* is assumed to be a uniform random variable. If this assumption is violated, the proportion of compliers cannot be calculated.

## References

Abrigo MR, Halliday T, Molina T (2019) Expanding health insurance for the elderly of the Philippines. IZA Discussion Paper

Aldrich DP, Yoshida T (2020) How Japan Stumbled into a Pandemic Miracle. Current History 119(818):217–221

American Hospital Association (2020) Hospitals and health systems face unprecedented financial pressures due to COVID-19. URL https://www.aha.org/guidesreports

Carneiro P, Løken KV, Salvanes KG (2015) A flying start? maternity leave benefits and long-run outcomes of children. Journal of Political Economy 123(2):365–412

Cornelissen T, Dustmann C, Raute A (2018) Who benefits from universal child care? estimating marginal returns to early child care attendance. Journal of Political Economy 126(6):2356–2409

GHC (2020) The third report: The number of patients with COVID-19 increased further, and the number of patients with cancer, cerebral infarction and heart failure also decreased. URL https://gemmed.ghc-j.com/?p=35059

Gruber J, Owings M (1996) Physician financial incentives and cesarean section delivery. The RAND Journal of Economics 27(1):99–123

Heckman JJ, Vytlacil E (2001) Policy-relevant treatment effects. American Economic Review 91(2):107–111

Heckman JJ, Vytlacil E (2005) Structural equations, treatment effects, and econometric policy evaluation 1. Econometrica 73(3):669–738

Heckman JJ, Vytlacil EJ (2007) Econometric evaluation of social programs, part ii: Using the marginal treatment effect to organize alternative econometric estimators to evaluate social programs, and to forecast their effects in new environments. Handbook of econometrics 6:4875–5143

Heckman JJ, Ichimura H, Todd PE (1997) Matching As An Econometric Evaluation Estimator: Evidence from Evaluating a Job Training Programme. The Review of Economic Studies 64(4):605–654

Ikegami N, Campbell JC (1995) Medical care in Japan. New England Journal of Medicine 333(19):1295–1299

Imbens GW, Angrist JD (1994) Identification and estimation of local average treatment effects. Econometrica 62(2):467–475

Japan Hospital Association (2020) Special survey on the current situation of hospital management. URL http://www.hospital.or.jp/pdf/06_20200527_01.pdf

Japan Times (2020) Hospitals in Japan cut bonuses as coronavirus drives them into red. URL https://www.japantimes.co.jp/news/2020/07/14/business/corporate-business/hospitals-japan-cut-staff-bonuses-coronavirus/

Kamhöfer DA, Schmitz H, Westphal M (2019) Heterogeneity in marginal non-monetary returns to higher education. Journal of the European Economic Association 17(1):205–244

Khan AA, AlRuthia Y, Balkhi B, Alghadeer SM, Temsah MH, Althunayyan SM, Alsofayan YM (2020) COVID-19 Survival and Cost in Saudi Arabia. International journal of environmental research and public health 17(20):7458

Kobayashi K (2020) Six propositions to prevent medical collapse. https://cigs.canon/article/20200925_5371.html

Kowalski AE (2016) Doing more when you’re running late: Applying marginal treatment effect methods to examine treatment effect heterogeneity in experiments. Tech. rep., National Bureau of Economic Research

Marbach M, Hangartner D (2020) Profiling compliers and noncompliers for instrumental-variable analysis. Political Analysis 28(3):435–444, DOI 10.1017/pan.2019.48

MHLW (2018) Survey on medical institutions. URL https://www.mhlw.go.jp/toukei/saikin/hw/iryosd/18/dl/02sisetu30.pdf

MHLW (2019) Current situation on the Type2-IDDH. URL https://www.mhlw.go.jp/bunya/kenkou/kekkaku-kansenshou15/02-02.html

MHLW (2020a) Prepared for a significant increase in the number of patients with COVID-19. URL https://www.mhlw.go.jp/content/000614594.pdf

MHLW (2020b) Securing beds for patients infected with the novel coronavirus (Request). URL https://www.mhlw.go.jp/content/10900000/000593853.pdf

Moynihan R, Johansson M, Maybee A, Lang E, Ĺegaŕe F (2020) COVID-19: an opportunity to reduce unnecessary healthcare. BMJ 370, DOI 10.1136/bmj.m2752, https://www.bmj.com/content/370/bmj.m2752.full.pdf

NHS England and NHS Improvement Website (2021) COVID-19 hospital activity. URL https://www.england.nhs.uk/statistics/statistical-work-areas/covid-19-hospital-activity/

OECD (2020) OECD health statistics 2020. URL http://www.oecd.org/els/health-systems/health-data.htm

Rodwin VG, Gusmano MK (2006) Growing Older in World Cities: New York, London, Paris, and Tokyo. Vanderbilt University Press

Sacchetto D, Raviolo M, Beltrando C, Tommasoni N (2020) COVID-19 surge capacity solutions: our experience of converting a concert hall into a temporary hospital for mild and moderate COVID-19 patients. Disaster Medicine and Public Health Preparedness p 1–10, DOI 10.1017/dmp.2020.412

Takaku R (2020) How is increased selectivity of medical school admissions associated with physicians’ career choice? a Japanese experience. Human Resources for Health 18:1–15

TMA (2020) Press conference on October 2020. https://www.tokyo.med.or.jp/press_conference/tmapc20201013

Tokyo Metropolitan Government (2018) Survey on medical institutions in Tokyo. URL https://www.fukushihoken.metro.tokyo.lg.jp/kiban/chosa_tokei/iryosisetsu/heisei29nen/29pdf.files/29-iryousisetu1.pdf

Tokyo Metropolitan Government (2019) Patient survey in Tokyo. URL https://www.metro.tokyo.lg.jp/tosei/hodohappyo/press/2019/11/11/documents/09.pdf

Yip WC (1998) Physician response to medicare fee reductions: changes in the volume of coronary artery bypass graft (CABG) surgeries in the medicare and private sectors. Journal of Health Economics 17(6):675 – 699

Zhu W, Wang Y, Xiao K, Zhang H, Tian Y, Clifford SP, Xu J, Huang J (2020) Establishing and managing a temporary coronavirus disease 2019 specialty hospital in Wuhan, China. Anesthesiology: The Journal of the American Society of Anesthesiologists 132(6):1339–1345

